# Community burden of diphtheria during the 2023–24 epidemic in Kano State, Nigeria: a population-based household survey

**DOI:** 10.64898/2026.04.10.26348327

**Authors:** Suleiman Hudu, Khadidja Uthman, Yves Katuala, Imam Wada Bello, Yanu Mbuyi, Dagemlidet Tesfaye Worku, Sedi Carvalho Mbelani, Ichola Ismael Adjaho, Etienne Gignoux, Cheick Oumar Doumbia, Franck Ale, Jonathan Polonsky

## Abstract

**Background:** Since late 2022, Nigeria has experienced one of its largest recorded diphtheria epidemics, centred on Kano State, where facility-based surveillance documented over 18,000 cases between February 2022 and April 2024. The true community burden remains unknown. We conducted a population-based household survey to estimate community attack rates, mortality, vaccination coverage, and determinants of infection and death.

**Methods and Findings:** We performed a retrospective household survey (September–October 2024) in four Local Government Areas (LGAs) using spatially randomised cluster sampling (78 clusters; recall from January 2023). Survey-weighted logistic regression adjusted for age group, sex, vaccination, LGA, socioeconomic quintile, and household size.

We enrolled 7,998 individuals from 1,068 households (cooperation rate 98.6%). Among 7,889 with known diphtheria status, the community attack rate was 1.1% (95% confidence interval [CI] [0.7,1.4]). The attack rate was 3.1 times (95% CI [2.1,4.1]) that notified at three Médecins Sans Frontières-supported sites over the same period. The case fatality ratio was 8.7% (95% CI [3.9,18.5]; 8/88) overall and 21.3% (95% CI [5.1,57.5]; 3/15) under five. Of 110 all-cause deaths, 62.5% occurred at home, as did 4 of 8 diphtheria deaths, all after seeking care. Care-seeking delays of ≥4 days were associated with higher case mortality (risk ratio 24.3, 95% CI [2.97,199]; p=0.004). Exploratory all-cause and diphtheria-specific mortality rate ratios (reported unvaccinated versus vaccinated) were 2.03 (95% CI [1.19,3.48]) and 1.00 (95% CI [0.21,4.80]). Routine immunisation coverage was 38.0%; 45.7% (95% CI [38.9,52.5]) of 3,287 children were reported as unvaccinated. The main limitation is that diphtheria, cause of death, and vaccination status were respondent-reported without laboratory or record confirmation.

**Conclusions:** Community diphtheria burden appeared to exceed facility surveillance estimates, and most all-cause deaths occurred at home. Delayed care-seeking was associated with diphtheria mortality, highlighting the need for improved community surveillance, decentralised care, and better-targeted vaccination.

**Author Summary:** *Why Was This Study Done?:* - Diphtheria has re-emerged as an epidemic disease across West and Central Africa, and facility-based surveillance, the main source of outbreak data, may substantially underestimate true community burden if many cases and deaths never reach a health facility. - During a large 2023-2024 epidemic in Kano State, Nigeria, we wanted to know how much diphtheria illness and death facility surveillance was missing, and what factors most affected whether someone survived.

*What Did The Researchers Do And Find?:*

- We conducted a population-based household survey of nearly 8,000 people across four high-burden areas of Kano State, asking about diphtheria illness, care-seeking, vaccination, and deaths during the epidemic.
- From January 2023 to October 2024, the community attack rate was approximately three times the rate of cases recorded at three treatment sites supported by Médecins Sans Frontières in the same areas (approximately 1.9 times when only cases reported as diagnosed by a health worker were counted). These sites notified most (87%) of the suspected cases reported for Kano State up to September 2024, so the gap between cases in the community and cases seen in all health facilities may be somewhat smaller. Approximately two thirds of deaths from any cause occurred at home, as did 4 of the 8 deaths from diphtheria.
- Delay in seeking care was associated with death among people with diphtheria. Routine and campaign vaccination coverage were both incomplete, and the vaccination campaign reached children who were already vaccinated more often than children who had not been routinely vaccinated.

*What Do These Findings Mean?:*

- Facility-based surveillance alone may underestimate the toll of a diphtheria epidemic, and community-based approaches are needed to capture the full burden.
- Reducing delays to treatment, for example through decentralised care such as the home-based care model evaluated in a companion study, and redesigning vaccination campaigns to specifically reach previously unvaccinated children rather than re-dosing those already covered, could meaningfully reduce morbidity and mortality in future epidemics.
- These findings rest on what household members remembered and reported: diphtheria, causes of death, and vaccination were not confirmed by laboratory tests or medical records, and the timing of vaccination relative to illness was rarely recorded. Only 8 diphtheria deaths occurred, so estimates about death are imprecise, and the survey covered four heavily affected areas of Kano State, so the results may not apply elsewhere.

## Introduction

Diphtheria, caused by toxigenic *Corynebacterium diphtheriae*, is a vaccine-preventable infection in which systemic toxin dissemination can produce life-threatening myocarditis and polyneuropathy [1,2]. Global incidence declined after the introduction of the Expanded Programme on Immunization (EPI) in 1974, but epidemics continue where vaccination coverage has declined or health systems have been disrupted [3–5], as illustrated by recent epidemics in Yemen [6], Venezuela [7], and among migrant populations in Bangladesh [8] and Europe [9].

Since late 2022, West and Central Africa has experienced its worst diphtheria epidemic in decades, with nine countries reporting epidemics [10]. Nigeria has been by far the most affected country, reporting over 25,000 confirmed cases and more than 1,300 deaths nationally as of January 2026 [11]. Within Nigeria, Kano State has been the epicentre, accounting for approximately two thirds of all confirmed cases and deaths [12,13]. An analysis of 18,320 facility-based cases from the Kano State surveillance system documented a case fatality ratio (CFR) of 4.5% [13].

However, facility-based surveillance captures only patients who present to health services. Where the population faces barriers to treatment seeking at health facilities, a substantial proportion of cases and deaths may occur in the community without being counted [14,15]. Understanding this gap is essential for designing appropriate epidemic responses.

Médecins Sans Frontières (MSF), in collaboration with the Kano State Ministry of Health, supported a comprehensive diphtheria response from January 2023, including facility-based diphtheria treatment centres (DTCs), a home-based care programme for mild cases [16], vaccination campaigns, and community engagement. Despite these efforts, the extent of community transmission, and of mortality not captured by facility-based surveillance, remained uncertain.

We therefore conducted a population-based retrospective household survey in four of the most affected Local Government Areas (LGAs) of Kano State. We asked how the community attack rate (AR) and CFR compared with facility-based estimates, what the all-cause and diphtheria-specific mortality rates were, what proportion of children were vaccinated and whether reactive campaigns reached children without routine vaccination, and which factors were associated with infection and death.

## Methods

### Study design and setting

We performed a retrospective, population-based household survey in four LGAs of Kano State (Dawakin Tofa, Fagge, Nassarawa, and Ungogo) selected on the basis of their high case burden during the epidemic. From 1 January 2023 to 25 October 2024, the period of the survey recall, 9,864 residents of these four LGAs (348 per 100,000) with symptom onset in that period were notified as diphtheria cases at three MSF-supported treatment sites (Murtala Muhammad Specialist Hospital, the Infectious Diseases Hospital, and Waziri Shehu Gidado General Hospital) (MSF line list; aggregated counts in S2 Table). By 5 September 2024, these sites had notified 19,459 cases, equivalent to 86.7% of the 22,457 suspected cases reported for Kano State to the Nigeria Centre for Disease Control and Prevention [17]. Among the four LGAs, the notified rate was highest in Fagge (601 per 100,000), and the CFR among notified cases was highest in Dawakin Tofa (9.8%, 47 deaths among 481 cases; 11.1% among the 422 cases with a recorded outcome). Data collection took place between 17 September and 25 October 2024. The recall period extended from 1 January 2023, the approximate start of the epidemic, to the date of interview, providing an observation window of approximately 21 months. The study is reported following the Consensus-Based Checklist for Reporting of Survey Studies (CROSS) [18] (S1 Checklist).

### Protocol and analysis plan

The study followed a protocol (version 2024-07-08; S2 File) that the MSF Ethics Review Board approved on 9 July 2024 (reference 2437) and the Kano State Health Research Ethics Committee approved on 13 June 2024 (reference SHREC/2024/5164), before data collection began. The protocol also covered a matched cohort study of home-based care, reported separately [16]. For the household survey, it specified the sampling design and sample size; estimation of the incidence, the CFR, and the crude, under-five, and diphtheria-specific mortality rates with design-adjusted 95% confidence intervals; description of vaccination coverage and care-seeking; and Pearson χ² and t-tests for comparisons. It listed the estimation of vaccine effectiveness as a secondary objective; this was not estimated, because the timing of vaccination relative to illness was rarely recorded. The survey departed from the protocol in its timing (September– October 2024, planned for July–August 2024) and cluster design (78 clusters of approximately 15 households, planned as 90 clusters of 10), and the planned telephone verbal autopsy interviews were not conducted. Analyses not specified in the protocol, including those added in response to peer review, are identified as post hoc in the sections below.

### Sampling

We used a two-stage spatially randomised sampling design. Sixty-five random global positioning system (GPS) coordinates were generated across the four LGAs using the Epicentre GeoSampler application [19], and a further 13 coordinates in Ungogo were drawn from the reserve list of replacement points, giving 78 clusters and 1,068 consenting households. Eight clusters included households in two LGAs, so the analysis treated 86 LGA-specific clusters as the primary sampling units (Dawakin Tofa 11, Fagge 9, Nassarawa 24, Ungogo 42). Field enumerators enrolled the nearest 15 households to each coordinate to form clusters. The sample size of 899 households was calculated to estimate the crude mortality rate with a precision of ±0.08 per 10,000 person-days (95% confidence, design effect 1.5, 10% non-response). The household cooperation rate (American Association for Public Opinion Research [AAPOR] Cooperation Rate 1) was calculated as the number of households that consented to participate divided by the number of eligible households contacted.

### Participants

All individuals who had resided in the study areas at any point during the recall period were eligible. A household was defined as a group of individuals under the responsibility of one household head who customarily share meals and living space. Households were eligible if at least one adult member had resided continuously in the area throughout the recall period and could report on demographic events and illness episodes for all members. Household rosters recorded, for each member, whether they were present at interview, had left the household during the recall period, or had died, so that household composition could be reconstructed for the full recall window rather than assessed only at the point of interview.

### Data collection

Trained enumerators fluent in Hausa and English conducted interviews using a standardised questionnaire administered on tablets (KoboCollect) [20]. Teams received structured training covering ethical conduct, informed consent, interviewing techniques, and case definitions. The questionnaire covered household composition, demographics, vaccination history (with card verification where available), diphtheria-like illness episodes, health-seeking behaviour, outcomes, sequelae, perceptions of vaccination, and household assets for socioeconomic status derivation. An events calendar assisted respondents in locating events within the recall period. Supervisors conducted daily debriefings and random spot-checks of completed questionnaires.

### Definitions

A *diphtheria case* was defined as any household member reported by the respondent as having had suspected diphtheria during the recall period. This single-item approach was adopted for feasibility in a large household survey administered by lay interviewers during an actively publicised epidemic response, when community awareness of diphtheria’s clinical presentation was high. Respondents reporting a suspected case were asked a structured follow-up symptom checklist (including sore throat, difficulty swallowing, visible pharyngeal membrane, and neck swelling) and whether the case had been diagnosed by a health professional. These items were used to characterise reported cases and, in a post hoc sensitivity analysis, to define a restricted case definition (cases reported by the respondent as having been diagnosed by a health professional).

*Deaths* included all deaths reported among household members during the recall period. For every death, respondents were asked where the person died and whether health care had been sought in the 2 weeks before death. Respondents were asked a general cause-of-death question (a fixed list of leading causes, including malaria/febrile illness, diarrhoeal disease, malnutrition, and other categories) and, separately, whether diphtheria was believed to be the cause; the latter took precedence when reported. Respondents who reported a diphtheria death were asked a structured follow-up module covering symptoms, health-seeking behaviour, and treatment, mirroring the module used for living cases. No clinical, laboratory, or verbal-autopsy confirmation of cause of death was performed; cause of death was ascertained entirely from respondent report at the time of the household interview.

*Vaccination* was defined as reported receipt of at least one dose of a diphtheria-containing vaccine through routine immunisation or supplementary campaigns, at any time before or during the recall period, verified from vaccination cards where available; a completed three-dose primary series was not required. For sensitivity analyses, vaccination was additionally classified by source of evidence (card-confirmed or self-reported) and by route (routine only, campaign only, or both).

### Outcomes

Primary outcomes were the community AR, CFR, crude mortality rate (CMR), under-five mortality rate (U5MR), diphtheria-specific mortality rate, and intra-household secondary attack rate (SAR). Secondary outcomes were vaccination coverage (routine, campaign, and combined) and factors associated with infection and death.

### Statistical analysis

All analyses were conducted in R (version 4.5.2) using the *survey* package (version 4.5) to account for cluster design. We used design-based survey estimation to obtain population-averaged estimates, accounting for the two-stage cluster design, LGA stratification, and sampling weights; variances were estimated by Taylor series linearisation.

Proportions were expressed with design-adjusted 95% confidence intervals (95% CIs); CFRs are reported with logit-transformed survey CIs (post hoc), which remain within 0– 100% when deaths are few. Attack rates were calculated as the proportion of the survey population experiencing diphtheria. To quantify the gap between community and facility-based estimates, we computed (post hoc) the ratio of the survey-derived AR to the AR among cases notified at three MSF-supported treatment sites (the MSF-site AR, 348 per 100,000). The MSF-site AR was calculated from the MSF line list (aggregated counts in S2 Table) as the number of residents of the four LGAs notified as diphtheria cases at these sites with symptom onset from 1 January 2023 to 25 October 2024, the survey recall period (9,864; notification date was used for 11 cases with no onset date), divided by the population of the four LGAs (2,837,111, from WorldPop and National Population Commission estimates). The 95% CI of the ratio was derived by dividing the survey AR confidence bounds by the MSF-site AR, treated as a fixed denominator, and reflects survey sampling uncertainty only. The MSF-site AR excludes cases notified at other facilities. Crude, under-five, and diphtheria-specific mortality rates were calculated as deaths per 10,000 person-days, using person-time derived from household composition and reported entry and exit events. The intra-household SAR (post hoc) was calculated as the proportion of susceptible household members in affected households who developed diphtheria following the index case. Socioeconomic status was derived by principal component analysis of household-level asset indicators and classified into quintiles (post hoc). To assess campaign targeting (post hoc), routine EPI and campaign vaccination status were distinguished using card evidence and campaign phase dates. Records with ambiguous source attribution were treated as missing. We cross-tabulated routine EPI status against campaign vaccination among campaign-eligible children (aged 6 months to 15 years) with a classified vaccination source, and compared the proportions vaccinated in the campaign with a design-based Rao–Scott F test. This complete-case analysis included 3,287 of the 3,536 campaign-eligible children and excluded 249: vaccination status missing (54) and vaccination source unclassifiable (195). To explore geographic patterns (post hoc), we produced kernel-smoothed maps of diphtheria AR, U5MR, and vaccination coverage (routine EPI, campaign, and either), following the cluster-level empirical-Bayes shrinkage and kernel-smoothing method used in a previous outbreak investigation [21]. Spatial association was tested using Spearman rank correlation on the raw (unshrunk) cluster-level estimates.

Characteristics of cases and non-cases were compared with Pearson χ² tests with the Rao–Scott adjustment, and ARs by sex with a design-based t-test. Survey-weighted logistic regression was used to examine factors associated with diphtheria (post hoc); crude and adjusted odds ratios (ORs) were estimated. The multivariable model included age group (0–5, 6–10, 11–15, ≥16 years), sex, reported vaccination status, LGA, socioeconomic quintile, and household size. Reported vaccination was compared (post hoc) by whether the household respondent believed that vaccination protects against disease, using survey-weighted logistic regression adjusted for age group, sex, and socioeconomic quintile.

Among diphtheria cases, associations of treatment delay, vaccination, age, and sex with death were examined. Because only eight diphtheria deaths occurred, a saturated multivariable model or age-adjusted estimation among cases was not possible, and these analyses were primarily univariable. We estimated (post hoc) the absolute change in the risk of death per additional day of treatment delay using a linear probability model, and compared the risk of death among cases presenting ≥4 versus <4 days after symptom onset as a survey-weighted risk ratio (quasi-Poisson regression with a log link). As a post hoc sensitivity analysis, this ratio was recalculated after excluding deaths with inconsistent dates (symptom onset after the reported date of death, or a reported care-seeking delay longer than the interval from onset to death).

The association between reported vaccination status and death was examined in an exploratory (post hoc) analysis, because the timing of vaccination relative to illness was rarely recorded. In the whole surveyed population, excluding individuals with missing vaccination status, we compared all-cause and diphtheria-attributed mortality between individuals reported as unvaccinated and those reported as vaccinated as ratios of survey-weighted mortality rates (deaths per person-day), and estimated an all-cause rate ratio adjusted for age group (under 5 years and 5 years or older) from a survey-weighted quasi-Poisson model with log person-time as an offset. Among cases with known vaccination status, CFRs were compared by vaccination status with a design-based Rao–Scott test. For each unadjusted ratio of rates (the all-cause and diphtheria-attributed ratios), the 95% CI and p-value were obtained on the log scale from a Wald test, using a delta-method standard error that treats the two groups’ rates as independent.

Sensitivity analyses (post hoc) of the association between vaccination and diphtheria compared individuals with card-confirmed vaccination with individuals reported as unvaccinated, and stratified by vaccination route (routine versus campaign). In a further sensitivity analysis, the primary case definition (respondent-reported suspected diphtheria) was restricted to cases reported by the respondent as diagnosed by a health professional, and the AR, its ratio to the MSF-site AR, the CFR, and the vaccination-diphtheria association were recalculated under this restricted definition.

### Ethics

The MSF Ethics Review Board approved the protocol (version 2024-07-08; ID 2437) on 9 July 2024, and the Kano State Health Research Ethics Committee approved the study (reference SHREC/2024/5164, letter dated 13 June 2024; NHREC registration number NHREC/17/03//2018). Written informed consent was obtained from each household head, using signature or, for illiterate participants, fingerprint; for illiterate household heads, an independent witness chosen by the participant and unconnected to the research team attested that the information was accurately explained and that consent was given freely. Data were anonymised and stored in accordance with data protection regulations.

## Results

### Study population

Of 1,083 households approached, 1,068 (98.6%) consented to participate, providing data on 7,998 individuals including 110 who had died from any cause during the recall period. Diphtheria status was known for 7,889 individuals (88 cases and 7,801 non-cases; Table 1) and not recorded for 109, who included 2 of the 110 deaths. Analyses of diphtheria (ARs, CFRs, and Tables 1–3) use the 7,889 with known status, less those missing a variable in each analysis (vaccination status was missing for 558 non-cases and 2 cases); all-cause mortality analyses use all 7,998 individuals. Percentages reported with a 95% CI are survey-weighted; counts, and percentages reported without a CI, are unweighted. The sample was 51.1% female, with a median age of 17 years (interquartile range [IQR] 8–30); children aged 15 years or younger constituted 47% of the sample.

**Table 1.** Characteristics of survey participants with known diphtheria status, by diphtheria status. Counts (n) are unweighted; percentages are survey-weighted and exclude missing values. The table includes the 7,889 of 7,998 surveyed individuals whose diphtheria status was recorded; the 109 without recorded status include 2 of the 110 deaths. Vaccinated: reported receipt of at least one dose of diphtheria-containing vaccine; status missing for 558 individuals without diphtheria and 2 with diphtheria. p-values are from design-based Rao–Scott F tests. LGA, Local Government Area.

| Characteristic | No diphtheria<br>(N=7,801) | Diphtheria (N=88) | p-value |
| --- | --- | --- | --- |
| **LGA** |  |  | 0.2 |
| Dawakin Tofa | 874 (16%) | 14 (24%) |  |
| Fagge | 615 (13%) | 4 (7.6%) |  |
| Nassarawa | 2,260 (33%) | 17 (23%) |  |
| Ungogo | 4,052 (38%) | 53 (46%) |  |
| **Age group** |  |  | <0.001 |
| 0–5 years | 1,380 (17%) | 18 (19%) |  |
| 6–10 years | 1,247 (16%) | 25 (25%) |  |
| 11–15 years | 1,057 (14%) | 19 (25%) |  |
| ≥16 years | 4,117 (53%) | 26 (30%) |  |
| **Sex** |  |  | 0.071 |
| Female | 3,980 (51%) | 52 (61%) |  |
| Male | 3,821 (49%) | 36 (39%) |  |
| **Vaccinated** |  |  | 0.006 |
| No | 4,018 (57%) | 33 (38%) |  |
| Yes | 3,225 (43%) | 53 (62%) |  |
| **Outcome** |  |  | <0.001 |
| Alive | 7,701 (99%) | 80 (91%) |  |
| Died | 100 (1.2%) | 8 (8.7%) |  |

**Table 2.** Factors associated with diphtheria: crude and adjusted odds ratios from survey-weighted logistic regression. OR, odds ratio; CI, confidence interval; LGA, Local Government Area; SES, socioeconomic status; ref, reference category. Survey-weighted logistic regression with cluster-adjusted standard errors, among individuals with known diphtheria status and complete data for all covariates (multivariable model n=7,294; crude vaccination model n=7,329). Crude ORs are from models with one factor each; adjusted ORs are from one model including LGA, age group, sex, reported vaccination status, SES quintile (per quintile), and household size (per member).

| Factor | Crude OR<br>[95% CI] | p | Adjusted OR<br>[95% CI] | p |
| --- | --- | --- | --- | --- |
| **LGA (ref: Dawakin Tofa)** |  |  |  |  |
| Fagge | 0.41<br>[0.15,1.13] | 0.085 | 0.22<br>[0.08,0.59] | 0.003 |
| Nassarawa | 0.47<br>[0.17,1.31] | 0.147 | 0.28<br>[0.11,0.75] | 0.012 |
| Ungogo | 0.82<br>[0.33,2.02] | 0.657 | 0.56<br>[0.23,1.34] | 0.190 |
| **Age group (ref: ≥16 years)** |  |  |  |  |
| 0–5 years | 1.97<br>[1.05,3.71] | 0.035 | 1.34<br>[0.65,2.73] | 0.421 |
| 6–10 years | 2.84<br>[1.63,4.95] | <0.001 | 2.09<br>[1.12,3.90] | 0.021 |
| 11–15 years | 3.30<br>[1.76,6.17] | <0.001 | 2.79<br>[1.47,5.27] | 0.002 |
| **Sex (ref: male)** |  |  |  |  |
| Female | 1.51<br>[0.96,2.37] | 0.072 | 1.51<br>[0.96,2.36] | 0.072 |
| **Vaccinated** | 2.11<br>[1.23,3.64] | 0.008 | 1.91<br>[1.03,3.56] | 0.041 |
| **SES quintile (per quintile)** | 0.90<br>[0.73,1.12] | 0.354 | 0.75<br>[0.61,0.91] | 0.005 |
| **Household size (per member)** | 0.96<br>[0.92,1.00] | 0.067 | 0.94<br>[0.89,1.00] | 0.040 |

**Table 3.** Sensitivity analyses for the association between reported vaccination and diphtheria. OR, odds ratio; CI, confidence interval. Crude ORs from survey-weighted logistic regression, all ages, among individuals with known diphtheria and vaccination status. The card-confirmed analysis compares individuals reported as vaccinated with a card (a card seen, a card reported lost, or a dose reported at a DTC, outpatient department, or contact clinic) with individuals reported as unvaccinated, and excludes individuals reported as vaccinated without a card; cards reported lost and clinic-reported doses are recalled, not seen. The routine and campaign analyses compare individuals vaccinated by each route with individuals reported as unvaccinated; route is as reported by the caregiver: routine vaccination is a card seen, a card reported lost, or a dose reported at a DTC, outpatient department, or contact clinic, without a campaign dose reported, so these rows include recalled vaccination; campaign vaccination is any campaign dose reported.

| Analysis | OR | 95% CI | p |
| --- | --- | --- | --- |
| Full sample: reported vaccinated vs reported unvaccinated | 2.11 | [1.23,3.64] | 0.008 |
| Card-confirmed vaccination vs reported unvaccinated | 2.02 | [1.11,3.68] | 0.022 |
| Routine vaccination only vs reported unvaccinated | 2.01 | [1.01,4.01] | 0.046 |
| Any campaign vaccination vs reported unvaccinated | 2.69 | [1.42,5.10] | 0.003 |

### Community attack rates and comparison with facility notifications

Of the 7,889 individuals with known diphtheria status, 88 were reported to have experienced diphtheria, yielding a crude community AR of 1.1% (95% CI [0.7,1.4]) and an incidence rate of 6.5 cases per 1,000 person-years (95% CI [4.4,8.5]) (Table 1, Fig 1). The survey AR was 3.1 times (95% CI [2.1,4.1]) the MSF-site AR of 0.348% (348 per 100,000). Every reported case also endorsed at least one of the four core symptoms on the follow-up checklist (88/88). Restricting to the subset of cases reported by the respondent as having been diagnosed by a health professional (58/88, 65.9%) gave a lower attack rate (0.67%, 95% CI [0.44,0.91]) than the primary case definition (1.1%), and a higher CFR (11.3%, 95% CI [4.3,26.6]; 6/58 vs 8.7%); the positive association between vaccination and diphtheria persisted under this restricted definition (OR 2.32, 95% CI [1.07,5.02]; p=0.034), similar in magnitude to the primary full-sample estimate. Under the restricted definition, the survey AR was 1.9 times (95% CI [1.3,2.6]) the MSF-site AR.

**Fig 1.**
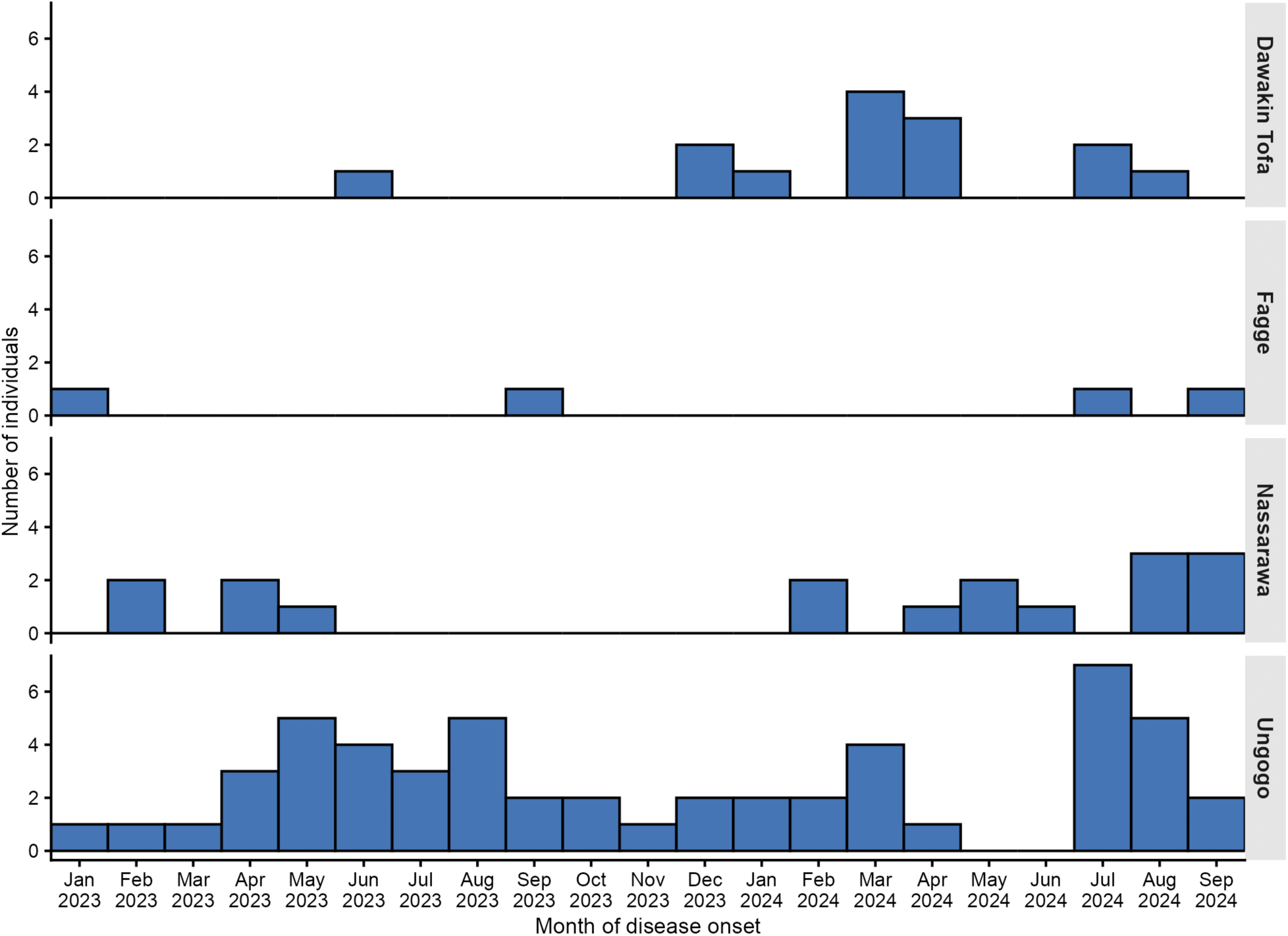
Epidemic curve of reported diphtheria cases by month of onset, household survey, Kano State, 2023–24.

ARs were highest among school-aged children: 2.0% (95% CI [0.9,3.1]) among those aged 11–15 years and 1.7% (95% CI [0.9,2.6]) among those aged 6–10 years. Adults (≥16 years) had the lowest AR (0.6%, 95% CI [0.4,0.9]). ARs were 1.3% (95% CI [0.8,1.8]) among females and 0.9% (95% CI [0.5,1.2]) among males (p=0.089, design-based t-test).

Among 63 households with at least one diphtheria case, 17 (27%) reported two or more cases, corresponding to an intra-household SAR of 5.2% (95% CI [2.1,8.4]).

### Factors associated with diphtheria

In multivariable analysis, school-aged children had higher odds of diphtheria compared with adults: adjusted OR 2.09 (95% CI [1.12,3.90]; p=0.021) for ages 6–10 years and 2.79 (95% CI [1.47,5.27]; p=0.002) for ages 11–15 years (Table 2). For children aged 0–5 years, the adjusted OR was 1.34 (95% CI [0.65,2.73]; p=0.421). Sex was not independently associated with diphtheria after adjustment, but socioeconomic quintile (adjusted OR 0.75 per quintile, 95% CI [0.61,0.91]; p=0.005) and LGA were: children in Fagge (adjusted OR 0.22, 95% CI [0.08,0.59]; p=0.003) and Nassarawa (adjusted OR 0.28, 95% CI [0.11,0.75]; p=0.012) had lower odds of diphtheria than those in Dawakin Tofa, the reference LGA, while the adjusted OR for Ungogo was 0.56 (95% CI [0.23,1.34]; p=0.190).

Reported vaccination remained associated with higher odds of diphtheria in the adjusted model (adjusted OR 1.91, 95% CI [1.03,3.56]). This association is examined further in the sensitivity analyses below.

### Mortality

There were 110 deaths from any cause among 7,998 survey participants during the recall period. The crude mortality rate was 0.21 per 10,000 person-days (95% CI [0.16,0.27]) and the under-five mortality rate was 0.93 per 10,000 person-days (95% CI [0.62,1.25]). The leading reported cause of death was malaria and other febrile illness (26.0%, 95% CI [17.4,34.6]), followed by diarrhoeal disease (9.8%, 95% CI [3.0,16.6]) and diphtheria (7.2%, 95% CI [2.2,12.1]). The diphtheria-specific mortality rate was 0.015 per 10,000 person-days (95% CI [0.004,0.027]; 8 deaths).

Eight deaths occurred among 88 diphtheria cases, giving a CFR of 8.7% (95% CI [3.9,18.5]) overall and 21.3% (95% CI [5.1,57.5]; 3 deaths among 15 cases) among children younger than five years. The median interval from symptom onset to death was 6 days (IQR 4–12; 8 deaths). Four of these records had an inconsistent date or delay (symptom onset after the reported date of death in 1, and a reported care-seeking delay longer than the interval from onset to death in 3); excluding them, the median was 8 days.

### Health-seeking behaviour and treatment delay

Seventy-six of the 88 cases (83.6%, 95% CI [69.8,97.4]) sought medical care during their illness, with a median delay from symptom onset to first consultation of 2 days (IQR 1–4). Among the 12 cases who did not seek care, reasons recorded included perceived low severity (n=2), facility distance (n=1), and traditional-healer use (n=1); the remainder gave an unspecified reason (n=7) or did not know (n=1). Health-seeking behaviour was associated with survival; the risk of death increased by 2.4 percentage points (95% CI [0.7,4.0]; p=0.006) for each additional day of delay, and those who waited four days or more had a higher risk of death than those who presented within three days (risk ratio 24.3, 95% CI [2.97,199]; p=0.004; 7 of 21 cases who waited four days or more died, compared with 1 of 55 who presented sooner). Four of the 8 diphtheria deaths had an inconsistent date or delay (see Mortality); excluding these records, the risk ratio was 16.2 (95% CI [1.47,177]; p=0.024).

Of the 110 deaths from any cause, 62.5% (95% CI [52.0,73.0]) occurred at home, 31.8% (95% CI [21.4,42.2]) in a health facility, and 4.3% (95% CI [0.3,8.3]) in transit. Respondents reported that care had been sought in the 2 weeks before death for 72 of the 110 deaths (no for 27, not known for 3, and missing for 8), including 47 of the 69 deaths at home. All 8 diphtheria deaths had been preceded by care-seeking: 4 occurred at home (2 after admission to a health facility and 2 after medication was given to take at home), 3 in a health facility after admission, and 1 in transit after medication was given to take at home.

### Vaccination and mortality

In this exploratory analysis, vaccination was defined as reported receipt of at least one dose of diphtheria-containing vaccine, by any route, at any time (see Methods: Definitions); the timing of vaccination relative to illness or death was unknown.

Across the whole surveyed population, 71 deaths from any cause occurred over 2,489,831 person-days among individuals reported as unvaccinated and 26 over 1,998,564 person-days among those reported as vaccinated (all-cause mortality rate ratio, unvaccinated versus vaccinated, 2.03, 95% CI [1.19,3.48]; p=0.010); 13 deaths among individuals with missing vaccination status were excluded. With adjustment for age group (0–4 and ≥5 years) in a quasi-Poisson model with a person-time offset, the all-cause mortality rate ratio, unvaccinated versus vaccinated, was 3.12 (95% CI [1.86,5.24]). For deaths attributed to diphtheria (4 among unvaccinated and 3 among vaccinated individuals), the mortality rate ratio, unvaccinated versus vaccinated, was 1.00 (95% CI [0.21,4.80]; p=1.00). Among the 86 diphtheria cases with known vaccination status, the survey-weighted CFR was 11.9% (95% CI [4.1,29.9]; 4 deaths among 33 cases) in unvaccinated cases and 5.6% (95% CI [1.5,18.9]; 3 deaths among 53 cases) in vaccinated cases (p=0.34, design-based Rao–Scott test).

### Vaccination and diphtheria: sensitivity analyses

In the full sample, individuals who reported being vaccinated had higher odds of diphtheria than those who reported being unvaccinated (OR 2.11, 95% CI [1.23,3.64]; p=0.008; Table 3). This paradoxical association was examined through sensitivity analyses.

When restricted to individuals with card-confirmed vaccination, compared with those reported as unvaccinated, the elevated odds persisted with little attenuation (OR 2.02, 95% CI [1.11,3.68]; p=0.022). Stratified analysis by vaccination route showed that the elevated odds also persisted among those vaccinated through routine immunisation (OR 2.01, 95% CI [1.01,4.01]; p=0.046) and among campaign-vaccinated individuals (OR 2.69, 95% CI [1.42,5.10]; p=0.003).

Among the 53 vaccinated cases, 14 (26.4%) reported being vaccinated specifically at a diphtheria treatment centre, outpatient department, or contact clinic. Vaccination date was recorded for only 3 of the 88 cases; of these, one was vaccinated the day after symptom onset and two were vaccinated more than a year before onset.

### Vaccination coverage and campaign performance

Among 3,536 campaign-eligible children aged 6 months to 15 years, survey-weighted routine EPI coverage was 38.0% (95% CI [31.7,44.3]) among the 3,396 with known EPI status, and campaign coverage was 35.6% (95% CI [29.5,41.8]) among the 3,486 with known campaign status. Among the 3,287 children with sufficient evidence to classify vaccination source (249 [7.0%] were unclassifiable and excluded), 54.3% (95% CI [47.5,61.1]) had evidence of vaccination from at least one source, and 45.7% (95% CI [38.9,52.5]) reported no vaccination of any kind.

Of the 3,536 children, vaccination status was missing for 54 (1.5%), 1,482 (41.9%) were reported as unvaccinated, and 2,000 (56.6%) were reported as vaccinated. The 3,287 children with a classified vaccination source were included in the analysis (Table 4); the 249 excluded had missing vaccination status (54) or a vaccination source that could not be classified (195). Of these, 1,280 were reported as having received routine EPI vaccination: a card was seen for 197, a card was reported lost for 894, and a dose was reported at a diphtheria treatment centre (DTC), outpatient department, or contact clinic for 189. The other 2,007 comprised the 1,482 reported as unvaccinated and 525 reported as vaccinated in a campaign. The survey-weighted proportion reported as unvaccinated was 43.5% (95% CI [36.9,50.2]) of the 3,482 children with known status, which includes the 1,482 counted above.

**Table 4.** Routine EPI and reactive campaign vaccination among campaign-eligible children aged 6 months to 15 years with a classified vaccination source (n=3,287) EPI, Expanded Programme on Immunization; DTC, diphtheria treatment centre; CI, confidence interval. Counts are unweighted, with unweighted row percentages in parentheses; the survey-weighted estimate is stated separately below. Reported routine EPI vaccination: a routine dose on a card seen by the interviewer (197 children), a card reported lost (894), or a dose reported at a DTC, outpatient department, or contact clinic (189). No reported routine EPI vaccination: 1,482 children reported as unvaccinated, 439 reported as vaccinated without a card, and 86 whose card showed a campaign dose. Campaign vaccinated: a campaign dose reported by the caregiver or shown on a card. Campaign phases: March–April 2023, August–October 2023, and November 2023–January 2024. Excluded: 249 of the 3,536 campaign-eligible children (vaccination status missing, 54; vaccination source unclassifiable, 195). Survey-weighted, 25.1% (95% CI [17.5,32.8]) of children without reported routine EPI vaccination were campaign vaccinated. Association tested with a design-based Rao–Scott F test (F=22.28 on 1 and 81 degrees of freedom, p<0.001).

|  | Campaign vaccinated | Not campaign vaccinated | Total |
| --- | --- | --- | --- |
| **Reported routine EPI vaccination** | 742 (58.0%) | 538 (42.0%) | 1,280 |
| **No reported routine EPI vaccination** | 525 (26.2%) | 1,482 (73.8%) | 2,007 |
| **Total** | 1,267 (38.5%) | 2,020 (61.5%) | 3,287 |

Campaign doses were received at a higher rate by children with prior reported routine EPI vaccination than by those without (742/1,280 [58.0%] and 525/2,007 [26.2%] respectively; design-based Rao–Scott F test, p<0.001; Table 4, N=3,287). Among children with confirmed campaign vaccination, 59.0% (95% CI [47.8,70.2]) had prior reported routine EPI vaccination. Among children without reported routine EPI vaccination, 25.1% (95% CI [17.5,32.8]) received a campaign dose.

Routine EPI and campaign coverage were positively correlated across the 78 clusters (Spearman rho 0.46, p<0.001; Fig 2). Kernel-smoothed maps of vaccination coverage, diphtheria attack rate, and mortality (S1 Fig) showed apparent visual overlap between areas of lower coverage and higher disease burden; formal cluster-level testing, however, found no association between vaccination coverage (routine EPI, any evidence of vaccination, or campaign vaccination) and the diphtheria attack rate, all-cause mortality rate, or under-five mortality rate (Spearman rho −0.01 to 0.16, all p>0.10).

**Fig 2.**
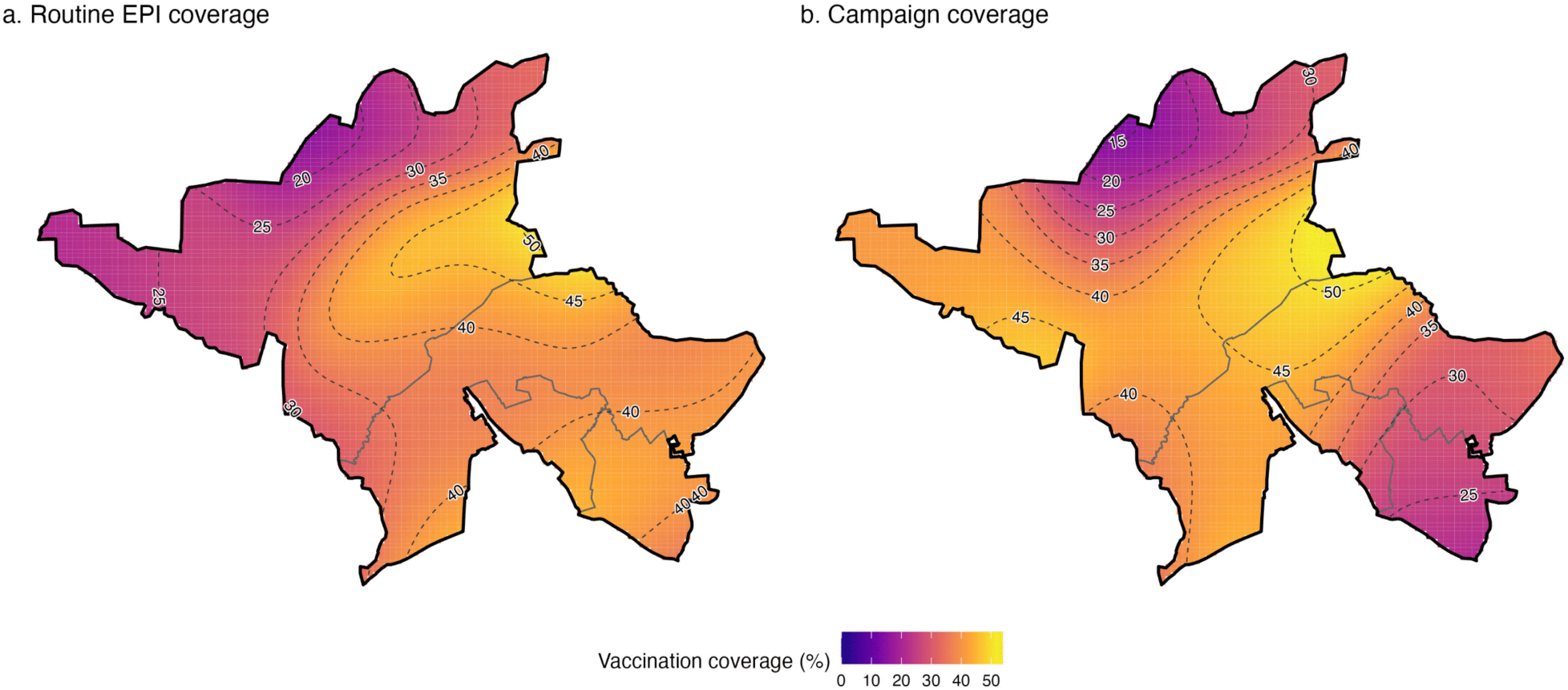
Kernel-smoothed maps of routine EPI (a) and reactive campaign (b) vaccination coverage among campaign-eligible children (6 months to 15 years), Kano State, 2023– 24. Administrative ward boundaries: GRID3 Nigeria Operational Ward Boundaries (eHealth Africa and Proxy Logics, 2020), Geo-Referenced Infrastructure and Demographic Data for Development (GRID3), https://data.grid3.org/datasets/GRID3::grid3-nga-operational-wards-v4-0/, licensed under CC BY 4.0 (https://creativecommons.org/licenses/by/4.0). EPI, Expanded Programme on Immunization.

Reported vaccination was more common among individuals in households whose respondent believed that vaccination protects against disease (3,225 of 7,066, 45.6%) than among those in households whose respondent did not (59 of 285, 20.7%); the adjusted odds ratio (OR) was 3.69 (95% CI [1.85,7.39]; p<0.001), adjusted for age group, sex, and socioeconomic quintile.

## Discussion

In this population-based household survey of a large diphtheria epidemic, the community attack rate was approximately three times the rate of cases notified at three MSF-supported treatment sites in the same areas over the same period, nearly one in ten cases died, and approximately two thirds of all deaths (any cause) occurred at home. These findings have implications for how epidemics are measured and managed across a region where at least eight countries remain affected [10,22].

Over the survey recall period, the community AR was approximately three times the AR among cases notified at three MSF-supported treatment sites in the same LGAs (1.9 times when restricted to cases reported as diagnosed by a health professional). In a setting where treatment capacity was overwhelmed [13,16], more than 16% of cases did not seek care, and most deaths from any cause occurred outside a health facility, passive surveillance is likely to have missed part of both morbidity and mortality, although care had been sought before most deaths, including all eight diphtheria deaths. A similar pattern has been described for measles and cholera in sub-Saharan Africa [23,24]. The three MSF-supported sites accounted for most cases reported for Kano State (86.7% of the suspected cases reported to NCDC by September 2024), but cases treated at other centres are not included, so the ratio may overstate the difference between community and facility counts. After MSF handed over two of the three sites in March 2024, only one site contributed to the line list, so completeness is likely to have been lower for later months; a completeness of 87% in the study LGAs over the whole recall period would put the ratio at approximately 2.7. The CI of the ratio reflects survey sampling uncertainty only. Reliance on facility-based case counts alone may therefore underestimate the scale of case-management, vaccination, and laboratory needs during large outbreaks.

Our community-based CFR of 8.7% was approximately twice the 4.5% from facility-based surveillance [13], although the confidence interval, based on eight deaths, includes the surveillance value. A higher community CFR, if confirmed, would be consistent with severe cases dying without being recorded by passive systems. Among children under five years, the CFR of 21.3% (3 deaths among 15 cases; 95% CI [5.1,57.5]) is compatible with pooled estimates of 20–30% for young unvaccinated children [25], but too imprecise to confirm them. School-aged children (6–15 years) bore the highest risk of infection, with odds more than double those of adults, consistent with congregate transmission in school settings and waning vaccine-derived immunity in a population with low routine coverage.

Diphtheria ranked third among reported causes of death, behind malaria or other febrile illness and diarrhoeal disease. This is plausible for a severe epidemic occurring in a setting where those two conditions are already leading causes of mortality. Cause of death was based entirely on respondent report, however, and these rankings should therefore be interpreted cautiously.

Each additional day of delay before seeking care was associated with a 2.4 percentage-point increase in the risk of death, and those who waited four or more days had a much higher risk of death than those who presented within three days (risk ratio 24.3), although this rested on a small number of events (1 of 55 versus 7 of 21) and persisted after excluding four deaths with inconsistent dates. This finding agrees with the companion cohort study evaluating home-based care during this epidemic, which similarly found that each additional day of delay was associated with higher odds of death (adjusted OR 1.29 per day) [16], and is consistent with established toxin kinetics: diphtheria antitoxin is most effective when administered within 48 hours of symptom onset, and efficacy declines rapidly with delay [2,25]; the reasons cases gave for not seeking care were too sparse and unspecific, however, to weigh facility access against care-seeking attitudes with any confidence. During a large epidemic in which treatment capacity is constrained, decentralised care that brings treatment closer to communities may shorten the critical interval between symptom onset and treatment. The companion study found that home-based management of mild cases achieved outcomes comparable to facility-based treatment in terms of mortality, household transmission, and patient acceptability [16].

The positive association between reported vaccination and diphtheria persisted after adjustment and in analyses restricted to card-confirmed vaccination. Reverse causality is a likely explanation. Vaccination was routinely offered to suspected cases and contacts during the outbreak response, and 14 of 53 vaccinated cases reported vaccination at a DTC, outpatient department, or contact clinic. This association was strongest among campaign-vaccinated individuals, where overlap between case presentation and vaccine delivery was greatest. Because vaccination dates were rarely recorded, we could not formally account for vaccination timing; however, the available descriptive evidence, including care-contact vaccination among cases and at least one documented post-exposure dose, is consistent with this mechanism.

Other explanations remain possible. Confounding by measured covariates is unlikely to account for the association, which persisted after adjustment for age, sex, socioeconomic status, LGA, and household size (adjusted OR 1.91, 95% CI [1.03,3.56]). Preferential targeting of high-risk, under-vaccinated households also appears unlikely, since campaign doses were received more often by children with reported routine EPI vaccination than by children without it. Recall and social-desirability bias cannot be excluded, particularly if respondents who recalled a diphtheria episode were also more likely to recall or report vaccination.

These findings also have implications for outbreak-response vaccination. Doses given after exposure or symptom onset cannot prevent the illness episode that prompted vaccination, so evaluations conducted during active epidemics should distinguish vaccination timing relative to exposure rather than relying on vaccination status alone. The lower campaign reach among children without reported routine vaccination also suggests that reactive campaigns may fail to reach those most in need of first-time protection. Future responses therefore need to address both when doses are delivered and whom they reach.

Reported vaccination was associated with lower all-cause mortality in the surveyed population, more strongly after adjustment for age group, but not with lower diphtheria-attributed mortality, and only 7 of the 97 deaths with known vaccination status were attributed to diphtheria. This all-cause association therefore cannot be read as protection by diphtheria vaccination. Vaccination status was reported at interview and its timing relative to illness was rarely known, so people who died before or early in the campaign periods had less opportunity than survivors to be vaccinated, which would introduce survival bias, favouring lower mortality among the vaccinated. Reported vaccination may also mark better access to health services or other characteristics associated with survival. These explanations could not be separated with the data available, and the analysis is exploratory.

Routine EPI coverage remained low (survey-weighted 38.0% of the 3,396 campaign-eligible children with known routine EPI status), and 45.7% of the 3,287 children whose vaccination source could be classified were reported as unvaccinated. Campaign vaccination was more common among children with reported routine EPI vaccination (742 of 1,280, 58.0%) than among those without it (525 of 2,007, 26.2%; unweighted), suggesting that the reactive campaign had limited success in reaching children without routine vaccination. Future campaigns should therefore explicitly identify and target populations missed by routine immunisation [21]. The association between belief in vaccine efficacy and uptake highlights the importance of community engagement during campaign planning and implementation, particularly amid reductions in international immunisation funding [26,27].

This study has several limitations. The retrospective design and long recall period (≥631 days) introduced recall bias, particularly for symptom onset and vaccination history. Survivor bias may have led to underestimation of attack and mortality rates, as households that dissolved following deaths or that relocated during the epidemic could not be sampled. Household composition could also change within enrolled households during the recall period through migration, birth, or death. At least one member left in 153 of 1,068 enrolled households (14.3%; 252 departures in total), and 75 individuals joined an enrolled household (52 households, 4.9%). Neither the date of departure nor of arrival was captured, so these transitions could not be linked to a specific date and were instead apportioned across the recall period using an interval-midpoint approximation for person-time calculations, which may introduce some imprecision into person-time-based mortality rates.

Self-reported vaccination status introduces misclassification, as the sensitivity analyses demonstrate. Vaccination status was missing for 54 of the 3,536 campaign-eligible children and the vaccination source could not be classified for a further 195, so the campaign-targeting analysis included 3,287 children. Routine and campaign vaccination were both reported by the caregiver, and for children without a card seen the dose could not be confirmed as routine rather than campaign, so the difference in campaign reach may be overstated. Coverage estimates and the targeting comparison should be interpreted with caution, as children with missing or unresolved vaccination data may differ systematically from those with complete records. The apparent spatial overlap between vaccination coverage and disease burden on our maps was not supported by formal cluster-level correlation testing; kernel smoothing can visually exaggerate similarity between surfaces beyond what the sparser underlying cluster-level data show, so these maps should be read as descriptive rather than as evidence of a spatial association.

The case definition relied on clinical recall rather than laboratory confirmation, and some misclassification of diphtheria status is likely. Other illnesses capable of producing a similar clinical picture, including bacterial tonsillitis, previously implicated in the low positive predictive value of a similarly constructed clinical case definition during the Rohingya diphtheria epidemic [8], as well as streptococcal pharyngitis, viral upper respiratory infections, and mumps (which can cause neck swelling), could plausibly have contributed to some misclassification, in addition to the malaria/febrile illness and diarrhoeal disease already discussed above in relation to cause-of-death ranking. Separately, because case ascertainment relied on the respondent’s own recognition of a suspected diphtheria episode, individuals more aware of diphtheria vaccination, whether through having been vaccinated themselves or through greater engagement with the outbreak response, may also have been more likely to correctly recognise and report an episode as diphtheria, a distinct differential-awareness mechanism from the recall and social-desirability biases already discussed above.

Case status, treatment-seeking, care sought before death, place of death, and cause of death were all based on respondent report; no linkage to medical records, facility registers, or death certificates was attempted. The health-professional-diagnosis figure reported above (Results) reflects respondent recall of that contact rather than independent verification, so the restricted case definition is not a confirmed case definition, and retrospective linkage to external records was not possible from the data analysed here.

Baseline characteristics of the 15 households that declined to participate were not captured, so a formal comparison between respondents and non-respondents was not possible. Households that could not be contacted are not included in the response rate, because the paper records of field visits were not available for analysis, so the response rate may overstate participation among all sampled households. Among households approached, the small number of refusals (1.4%) makes it unlikely that differential non-response materially biased the reported estimates.

Results from four high-burden LGAs may not generalise to areas with different epidemic intensity or health-service accessibility. The small number of diphtheria deaths (n=8) limited statistical power for the analysis of death among cases: models adjusting for additional covariates beyond age, sex, vaccination, and treatment delay became numerically unstable, consistent with quasi-complete separation, and were not pursued, and the reported treatment-delay estimates carry correspondingly wide confidence intervals. Four of the eight diphtheria deaths also had internally inconsistent dates (symptom onset after the reported date of death in one, and a reported care-seeking delay longer than the interval from onset to death in three), so the delay estimates and the interval from onset to death depend partly on uncertain recall, although the delay association remained after these records were excluded. The modest number of cases overall (n=88) similarly limited power in the multivariable model for infection risk.

The community burden of diphtheria during the 2023–24 epidemic in Kano State appeared to exceed what facility-based surveillance captured, and most deaths from any cause occurred at home, although care had been sought before most of them. Delayed care-seeking was associated with death among cases, routine EPI coverage remained low, and the reactive campaign reached children with reported routine vaccination more often than other children.

These findings support three priorities for future epidemic control: community-based surveillance to better capture disease burden; earlier access to treatment through decentralised care pathways, including home-based care for mild disease, which our companion cohort study found was not associated with worse outcomes [16]; and vaccination strategies that more effectively reach populations missed by routine and supplementary immunisation. As diphtheria continues to spread across West and Central Africa, strengthening these approaches will be important to reducing morbidity and mortality.

## Supporting information

S1 Checklist. CROSS reporting checklist

S1 Fig. Kernel-smoothed maps of vaccination coverage, attack rate and under-5 mortality

S1 File. Household survey questionnaire

S1 Table. Sensitivity analysis restricted to cases diagnosed by a health professional

S2 File. Study protocol (version 2024-07-08)

S2 Table. Diphtheria cases notified at three MSF-supported treatment sites

## Data Availability

Data used in this study are subject to restrictions arising from participant informed consent and General Data Protection Regulation (GDPR)-aligned privacy protections for identifying and potentially stigmatising health information, and cannot be deposited in a public repository. Anonymised individual-level data are available to qualified researchers upon request to the Epicentre Data Sharing Committee (https://epicentre.msf.org/en/our-projects/study-data-access-request), which reviews all requests independently of the study authors in accordance with Epicentre's Data Sharing Policy (https://epicentre.msf.org/sites/default/files/2020-02/Epicentre-Data%20Sharing%20Policy-EN-2020.pdf). Analysis code is available at https://github.com/jpolonsky/diphtheria-kano-2024, archived at Zenodo (https://doi.org/10.5281/zenodo.21907933).

## Supporting information

S1 Checklist. Consensus-Based Checklist for Reporting of Survey Studies (CROSS).

S1 Table. Sensitivity analysis restricting the case definition to cases reported as diagnosed by a health professional.

S1 Fig. Kernel-smoothed maps of vaccination coverage (any evidence), diphtheria attack rate, and under-5 mortality rate. Administrative ward boundaries: GRID3 Nigeria Operational Ward Boundaries (eHealth Africa and Proxy Logics, 2020), Geo-Referenced Infrastructure and Demographic Data for Development (GRID3), https://data.grid3.org/datasets/GRID3::grid3-nga-operational-wards-v4-0/, licensed under CC BY 4.0 (https://creativecommons.org/licenses/by/4.0).

S2 Table. Diphtheria cases notified at three MSF-supported treatment sites in Kano State, 2023 to 2024: aggregated counts used as the facility comparator. MSF, Médecins Sans Frontières.

S1 File. Household survey questionnaire.

S2 File. Study protocol (version 2024-07-08), covering the household survey and the companion matched cohort study.

## Acknowledgements

We thank the participating communities and local leaders of Dawakin Tofa, Fagge, Nassarawa, and Ungogo for their cooperation and hospitality. We are grateful to the field investigators and data teams for their dedication, and to the MSF WaCA coordination for logistical and operational support. The study was conducted by Epicentre in collaboration with MSF WaCA and the Kano State Ministry of Health. We acknowledge the ethical oversight provided by the Kano State Health Research Ethics Committee and the MSF Ethics Review Board.

## Contributors

JP, EG, and FA conceptualised the study and designed the methodology. JP, SH, KU, and FA led field planning and oversaw data collection. YK, IWB, YM, DTW, SCM, IIA, and COD contributed to study design and protocol development, and to data collection and field coordination. JP conducted the formal analysis and wrote the original draft. All authors contributed to critical review and revision of the manuscript for important intellectual content, and approved the final version for submission. JP accepts overall responsibility for the work.

## Funding

The author(s) received no specific funding for this work.

## Declaration of interests

We declare no competing interests.

## Notes

### Competing Interest Statement

The authors have declared no competing interest.

### Author Declarations

The Ethics Review Board of Médecins Sans Frontières gave ethical approval for this work (ID 2437, 9 July 2024). The Health Research Ethics Committee of the Kano State Ministry of Health, Nigeria, gave ethical approval for this work (reference SHREC/2024/5164, 13 June 2024; NHREC registration number NHREC/17/03//2018).

### Summary of Updates

This version is the revised manuscript resubmitted to PLOS Medicine on 22 October 2026 after peer review. The facility comparator is now the cases notified at the three MSF-supported treatment sites in the four study Local Government Areas over the whole recall period; the community attack rate was 3.1 times (95% CI 2.1 to 4.1) that rate, replacing 4.2 in version 2. Confidence intervals were recalculated with the 78 clusters as primary sampling units and without a finite population correction; the case fatality ratio is 8.7% (95% CI 3.9 to 18.5). The association between care-seeking delay and death is reported as a survey-weighted risk ratio (24.3, 95% CI 2.97 to 199). Vaccine effectiveness is no longer estimated; exploratory mortality rate ratios by reported vaccination status are reported instead. The study protocol, the facility notification counts, and the CROSS checklist are provided as supporting information.

