## Supplementary material for "Community burden of diphtheria during the 2023–24 epidemic in Kano State, Nigeria: a population-based household survey": S1 Checklist. CROSS reporting checklist

### S1 Checklist. Consensus-Based Checklist for Reporting of Survey Studies (CROSS).

| **Section** | **Item No** | **CROSS item description** | **Reported in manuscript (section, paragraph)** |
| --- | --- | --- | --- |
| Title and abstract | 1a | State the word “survey” along with a commonly used term in title or abstract to introduce the study's design. | Title (“...a population-based household survey”); Abstract, Methods and Findings (“retrospective household survey...spatially randomised cluster sampling”) |
|  | 1b | Provide an informative summary in the abstract, covering background, objectives, methods, findings/results, interpretation/discussion, and conclusions. | Abstract (Background / Methods and Findings / Conclusions) |
| Introduction | 2 | Provide a background about the rationale of study, what has been previously done, and why this survey is needed. | Introduction, paragraphs 1–4 (diphtheria and the West and Central African epidemic; limitations of facility-based surveillance; MSF response context) |
|  | 3 | Identify specific purposes, aims, goals, or objectives of the study. | Introduction, final paragraph (“We therefore conducted a population-based retrospective household survey...”) |
| Methods | 4 | Specify the study design in the methods section with a commonly used term (e.g., cross-sectional or longitudinal). | Methods, Study design and setting, paragraph 1 (“retrospective, population-based household survey”) |
|  | 5a | Describe the questionnaire (number of sections, number of questions, number and names of instruments used). | Methods, Data collection, paragraph 1; full instrument provided as S1 File |
|  | 5b | Describe all questionnaire instruments used to measure particular concepts. Report target population, validity/reliability information, scoring/classification procedure, and reference links. | Methods, Data collection and Definitions subsections (case definition, vaccination definition, cause-of-death ascertainment) |
|  | 5c | Provide information on pretesting of the questionnaire, if performed. | Not reported in the manuscript text (the questionnaire was piloted during interviewer training) |
|  | 5d | Questionnaire should be fully provided (in the article, or as appendices or as an online supplement). | S1 File (household survey questionnaire) |
|  | 6a | Describe the study population (background, locations, eligibility criteria for inclusion, exclusion criteria). | Methods, Participants, paragraph 1 |
|  | 6b | Describe the sampling techniques used. Specify the locations of sample participants whenever clustered sampling was applied. | Methods, Sampling, paragraph 1 (two-stage spatially randomised cluster design, 78 clusters across four LGAs) |
|  | 6c | Provide information on sample size, along with details of sample size calculation. | Methods, Sampling, paragraph 1 |
|  | 6d | Describe how representative the sample is of the study population, particularly for population-based surveys. | Methods, Sampling, paragraph 1 (household cooperation rate, AAPOR Cooperation Rate 1: consenting households as a proportion of households contacted); Results, Study population, paragraph 1 (98.6%, 1,068/1,083; 7,889 with recorded diphtheria status); Discussion, paragraph 15 (15 non-consenting households; non-contacts not included in the denominator) |
|  | 7a | Provide information on modes of questionnaire administration, including type/number of contacts and location. | Methods, Data collection, paragraph 1 (tablet-based interviews, KoboCollect, in-person household visits) |
|  | 7b | Provide information on the survey's time frame (recruitment, exposure, follow-up). | Methods, Study design and setting, paragraph 1 (data collection 17 September to 25 October 2024; recall period from 1 January 2023) |
|  | 7c | Provide information on the data entry process (error-minimisation for non-web-based surveys). | Methods, Data collection, paragraph 1 (electronic tablet-based data capture via KoboCollect; daily debriefings and spot-checks of completed questionnaires) |
|  | 8 | Describe any preparation process before conducting the survey (interviewer training, advertising). | Methods, Data collection, paragraph 1 (structured training covering ethical conduct, informed consent, interviewing technique, and case definitions) |
|  | 9a | Provide information on ethical approval (informed consent, IRB approval, Helsinki declaration, GCP declaration as appropriate). | Methods, Protocol and analysis plan, paragraph 1, and Ethics, paragraph 1 (MSF Ethics Review Board, reference 2437; Kano State Health Research Ethics Committee, reference SHREC/2024/5164, NHREC registration number NHREC/17/03//2018; written informed consent, including the witnessed fingerprint procedure for illiterate household heads) |
|  | 9b | Provide information about survey anonymity and confidentiality and mechanisms used to protect unauthorized access. | Methods, Ethics, paragraph 1 (data anonymised and stored in accordance with data protection regulations); Data sharing statement |
|  | 10a | Describe statistical methods and analytical approach. Report the statistical software used. | Methods, Statistical analysis, paragraph 1 (R version 4.5.2, survey package, design-based estimation with Taylor series linearisation) |
|  | 10b | Report any modification of variables used in the analysis, along with reference (if available). | Methods, Definitions (case, death, vaccination definitions); Statistical analysis (socioeconomic quintile derivation, campaign-eligibility restriction) |
|  | 10c | Report details about how missing data was handled (rate, mechanism, methods used). | Methods, Statistical analysis, paragraph 2 (complete-case analysis for campaign targeting); Results, Study population, paragraph 1 (diphtheria status not recorded for 109; vaccination status missing for 560) and Vaccination coverage and campaign performance, paragraph 2 (249 children excluded from Table 4: vaccination status missing, 54; vaccination source unclassifiable, 195); Table 1 and Table 4 footnotes; Discussion, paragraph 12. Multiple imputation was not used. |
|  | 10d | State how non-response error was addressed. | Methods, Sampling, paragraph 1 (household cooperation rate); Results, Study population, paragraph 1 (98.6%); Discussion, paragraph 15 (respondent and non-respondent comparison not possible; non-contacts not included in the denominator; 15 of 1,083 households declined) |
|  | 10e | For longitudinal surveys, state how loss to follow-up was addressed. | Not applicable, single cross-sectional retrospective interview, no follow-up |
|  | 10f | Indicate whether weighting of items or propensity scores were used to adjust for non-representativeness. | Methods, Statistical analysis, paragraph 1 (survey design weights applied with the survey package to account for the two-stage cluster design and stratification by LGA) |
|  | 10g | Describe any sensitivity analysis conducted. | Methods, Statistical analysis, paragraphs 4 and 6 (card-confirmed and route-stratified vaccination sensitivity analyses; restricted case definition; treatment-delay risk ratio excluding deaths with inconsistent dates); Results; S1 Table |
| Results | 11a | Report numbers of individuals at each stage of the study; consider a flow diagram. | Results, Study population, paragraph 1 (1,083 households approached, 1,068 consented, 7,998 individuals, 7,889 with recorded diphtheria status, 110 deaths) |
|  | 11b | Provide reasons for non-participation at each stage, if possible. | Not reported in the manuscript text; reasons for the 15 non-consenting households were not recorded (Discussion, paragraph 15) |
|  | 11c | Report response rate, present the definition or formula used. | Methods, Sampling, paragraph 1 (definition of the household cooperation rate); Abstract, Methods and Findings, and Results, Study population, paragraph 1 (98.6%, 1,068/1,083) |
|  | 11d | Define how unique visitors are determined (web-based surveys). | Not applicable, interviewer-administered household survey, not web-based |
|  | 12 | Provide characteristics of study participants, and information on potential confounders and assessed outcomes. | Table 1 (participant characteristics by diphtheria status: LGA, age group, sex, vaccination status, outcome) |
|  | 13a | Give unadjusted estimates and, if applicable, confounder-adjusted estimates with 95% CIs and p-values. | Table 2 (crude and adjusted odds ratios); Table 3 (crude sensitivity-analysis odds ratios); Table 4 (campaign-targeting cross-tabulation with Rao–Scott test); Results text throughout |
|  | 13b | For multivariable analysis, provide information on model building process, fit statistics, and assumptions. | Methods, Statistical analysis, paragraph 3 (a priori covariates specified: age group, sex, vaccination status, LGA, socioeconomic quintile, household size) |
|  | 13c | Provide details of any sensitivity analysis performed; for missing data, compare complete-case with imputed results if possible. | Methods, Statistical analysis, paragraphs 4 and 6 (sensitivity analyses); Results, Vaccination and diphtheria: sensitivity analyses; S1 Table. Complete-case analysis only; imputed results not compared. |
| Discussion | 14 | Discuss the limitations of the study, considering sources of potential bias and imprecision. | Discussion, paragraph 2 (comparator for the attack-rate ratio: three MSF-supported sites, 86.7% of NCDC suspected cases to September 2024, only one site contributing after March 2024) and paragraphs 11–16 (recall and survivor bias and household composition change; vaccination misclassification and missing or unclassifiable vaccination data; case-definition misclassification; respondent-report-only ascertainment; non-response; generalisability, statistical power, and date inconsistencies among diphtheria deaths) |
|  | 15 | Give a cautious overall interpretation of results, based on potential biases/imprecisions, and suggest areas for future research. | Discussion, paragraphs 1–10, and closing paragraphs 17 and 18 |
|  | 16 | Discuss the external validity of the results. | Discussion, paragraph 16 (results from four high-burden LGAs may not generalise to areas with different epidemic intensity or health-service accessibility) |
| Other sections | 17 | State whether any funding organization had a role in the survey's design, implementation, and analysis. | Funding section (“The author(s) received no specific funding for this work.”) |
|  | 18 | Declare any potential conflict of interest. | Declaration of interests (“We declare no competing interests.”) |
|  | 19 | Provide names of organizations/persons acknowledged, along with their contribution to the research. | Acknowledgements section |

Source checklist: Sharma A, Minh Duc NT, Luu Lam Thang T, Nam NH, Ng SJ, Abbas KS, et al. A Consensus-Based Checklist for Reporting of Survey Studies (CROSS). J Gen Intern Med. 2021;36(10):3179-3187. doi:10.1007/s11606-021-06737-1
