## Supplementary figures and images for "Community burden of diphtheria during the 2023–24 epidemic in Kano State, Nigeria: a population-based household survey"

### S1 Fig. Kernel-smoothed maps of vaccination coverage, attack rate and under-5 mortality

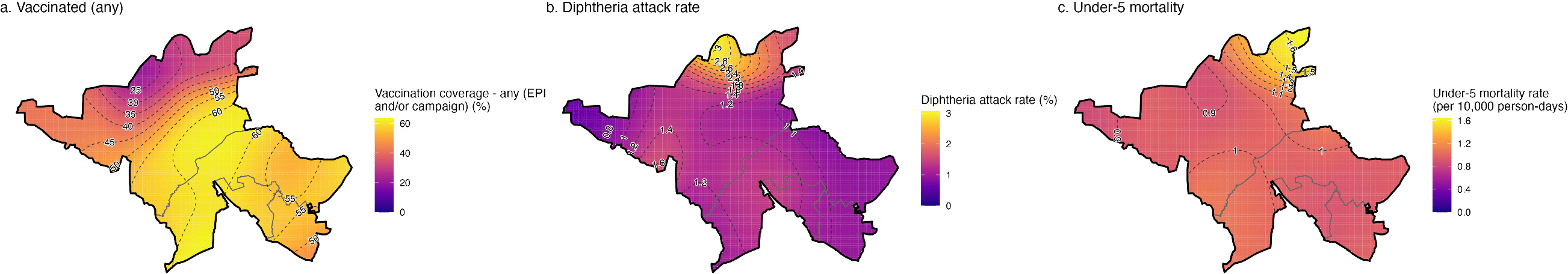
