## Supplementary material for "Community burden of diphtheria during the 2023–24 epidemic in Kano State, Nigeria: a population-based household survey": S1 File. Household survey questionnaire

| **Variable** | **Values** | **Comments** |
| --- | --- | --- |
| **CORE QUESTIONS** | | |
| Team ID | Team ID number |  |
| Date of interview | dd/mm/yyyy |  |
| Cluster ID | Numeric |  |
| Household ID | Numeric | Three-digit number |
| Written informed consent given by the head of the household | Y = Yes N = No |  |
| **HOUSEHOLD QUESTIONS** | | |
| GPS coordinates of household | Lon & Lat (and precision of point) |  |
| *Socio-economic status* |  |  |
| Socioeconomic status:  Household Effects | 0 = None of the following  1 = Radio  2 = Television  3 = Mobile Phone  4 = Computer  5 = Refrigerator  6 = Table  7 = Sofa  8 = Generator  9 = Fan  99 = DNK/NR | Multiple answers possible |
| Socioeconomic status:  Means of Transport | 0 = None of the following  1 = Bicycle  2 = Motorcycle/scooter  3 = Car/truck  4 = Keke napep  99 = DNK/NR | Multiple answers possible |
| Socioeconomic status:  Ownership of agricultural land | Y = Yes  N = No |  |
| Socioeconomic status:  Ownership of farm animals | 0 = None of the following  1 = Cows/bulls/other cattle  2 = Horses/Donkeys  3 = Goats/sheep  4 = Pigs  5 = Chickens/other poultry  6 = Camels  99 = DNK/NR | Multiple answers possible |
| Highest level of education of head of household | 0 = No education  1 = Some primary  2 = Completed primary  3 = Some secondary  4 = Completed secondary  5 = More than secondary |  |
| Highest level of education of female head of household (if there is a male head of household) | 0 = No education  1 = Some primary  2 = Completed primary  3 = Some secondary  4 = Completed secondary  5 = More than secondary |  |
| Main drinking water source | 1 = Surface  2 = Unimproved  3 = Improved  99 = DNK/NR | Surface water (rivers, lakes)  Unimproved water source (Unprotected dug well or unprotected spring)  Improved source (piped water, boreholes, tubewells, protected dug wells, protected springs, rainwater, packaged or delivered water) |
| Main type of toilet used by household members | 1 = Open defecation; 2 = Any unimproved; 3 = Improved 99 = DNK/NR | Open defecation (field, forest, nature)  Any unimproved facility (latrine without slab, bucket)  Any improved facility (toilets, pit latrines with slabs) |
| Handwashing facility with water & soap present at household | 0 = No  1 = Yes  99 = Unable to verify | Check in household |
| *Attitudes to vaccination* |  |  |
| Do you believe that vaccines can protect children from serious diseases? | 0 = No  1 = Yes |  |
| Do you think that most parents like you have their children vaccinated with all the recommended vaccines? | 0 = No  1 = Yes |  |
| Have you ever been reluctant or hesitated to get a vaccination for your child? | 0 = No  1 = Yes |  |
| Have you ever refused a vaccination for your child? | 0 = No  1 = Yes |  |
| If yes to questions 3 or 4, check which ones: |  |  |
| *Diphtheria vaccine* | 1 = Hesitated  2 = Refused |  |
| *DPT3 vaccine* | 1 = Hesitated  2 = Refused |  |
| *Polio vaccine* | 1 = Hesitated  2 = Refused |  |
| *Measles vaccine* | 1 = Hesitated  2 = Refused |  |
| *Meningococcal vaccine* | 1 = Hesitated  2 = Refused |  |
| *“Pentavalent” or other combination infant vaccine* | 1 = Hesitated  2 = Refused |  |
| *BCG vaccine* | 1 = Hesitated  2 = Refused |  |
| Reason for hesitation/refusal | 1 = Did not think it was needed  2 = Did not know where to get vaccination  3 = Did not know where to get good/reliable information  4 = Heard or read negative media  5 = Did not think the vaccine was effective  6 = Did not think the vaccine was safe/concerned about side effects  7 = Someone else told me that the vaccine was not safe  8 = Had a bad experience with previous vaccinator/health clinic  9 = Had a bad experience or reaction with previous vaccination  10 = Fear of needles  11 = Not possible to leave other work  12 = Religious reasons  13 = Refusal of male head of household  14 = Belief it can lead to infertility/sterilisation  15 = Other beliefs/traditional medicine  16 = Other (please specify): |  |
| Has distance ever prevented you from getting your child immunized? | 0 = No  1 = Yes |  |
| Has timing of clinic ever prevented you from getting your child immunized? | 0 = No  1 = Yes |  |
| Have costs ever prevented you from getting your child immunized? | 0 = No  1 = Yes |  |
| Are there other pressures that prevent you from getting your child immunized on time? | 0 = No  1 = Yes |  |
| Are there any reasons why children should not be vaccinated? | 0 = No  1 = Yes |  |
| Do you think it is difficult for some groups to get vaccinations? | 0 = No  1 = Yes |  |
| If yes, reason why: | 1 = They choose not to vaccinate  2 = They do not feel welcome at the health service  3 = Health services do not reach them |  |
| Have you ever received or heard negative information about vaccinations? | 0 = No  1 = Yes (specify) |  |
| If yes, did you still take your child to get vaccinated after? | 0 = No  1 = Yes |  |
| Which leaders in your community support vaccines for children? | 1 = Religious  2 = Political  3 = Teachers  4 = Health care workers 5 = Other (specify) | Multiple answers possible |
| Was your household visited by an MSF health promotion worker who provided information about diphtheria disease, treatment, and vaccination? | 0 = No  1 = Yes  99 = Do not recall |  |
| If yes, did this change how likely you would be to get vaccinations any children in your household? | 0 = No impact  1 = More likely  2 = Less likely  3 = Not relevant (no children in HH) |  |
| How many people are currently living in the household? | Numeric | Used to populate questionnaire (one for each ) |
| **INDIVIDUAL QUESTIONS (For each household member)** | | |
| *Household demography & mortality* |  |  |
| Household member order | Numeric |  |
| Sex | M = Male  F = Female |  |
| Age | Numeric (months/years) |  |
| Arrived during recall period? | 0 = No  1 = Yes |  |
| Born during recall period? | 0 = No  1 = Yes |  |
| Left during recall period | 0 = No  1 = Yes |  |
| Died during recall period | 0 = No  1 = Yes |  |
| If died, main cause of death? | 1 = Diphtheria  2 = Other respiratory infection  3 = Diarrhoea  4 = Malaria/Fever  5 = Malnutrition  6 = Measles  7 = Meningitis  8 = During pregnancy  9 = During/after delivery (<1 month)  10 = Trauma/accident  11 = Violence  12 = Other (specify)  99 = DNK |  |
| If diphtheria, was there a diagnosis by a health professional? | 0 = No  1 = Yes |  |
| If diphtheria, approximate date of disease onset? | Date |  |
| If died, place of death | 1 = At current location  2 = At the health centre  3 = At the hospital  4 = At DTC  5 = Other (specify)  99 = Don’t know |  |
| If died, did s/he seek health care in the 2 weeks before dying? | 0 = No  1 = Yes  99 = DNK |  |
| Place health care sought? | 1 = Home  2 = Health centre  3 = Diphtheria Treatment Centre (*specify*)  4 = Private hospital *(specify)*  5 = Public hospital (*specify*)  6 = Traditional healer  7 = Patient Medicine Vendor/Pharmacy  8 = Other place *(specify)*  99 = DNK |  |
| If not at a health facility, what was the main reason for not seeking care in a health structure? | 1 = No money/consultation too expensive  2 = Too sick to seek care  3 = Not sick enough to seek care 4 = Health facility too far away 5 = Went to traditional healer  6 = Did not have time/too busy to go 7 = No confidence in health system  8 = Security problem 9 = Care was refused at facility  10 = Refusal of male head of household 11 = Died suddenly before visit could be arranged  12 = Other reason *(specify)* 99 = DNK |  |
| *Diphtheria vaccination status* |  |  |
| Have you ever received diphtheria vaccination? | 0 = No  1 = Yes |  |
| If yes: Number of doses according to vaccination card | 0 = No card  1 = Yes (1 dose)  2 = Yes (2 doses)  3 = Yes (3 or more doses) |  |
| If card:  Date of vaccination (most recent dose) | (dd/mm/yyyy) |  |
| If no card:  Approximate vaccination date | (MM YYYY) |  |
| Is this individual aged 16 or under? | 0 = No  1 = Yes |  |
| If yes, did they receive the vaccine provided during the campaign? | 0 = No  1 = Yes (card observed)  2 = Yes (card not observed) | Specify months of campaign in question |
| If no, reason why not vaccinated | 1 = Did not think it was needed  2 = Did not know where to get vaccination  3 = Did not know where to get good/reliable information  4 = Heard or read negative media  5 = Did not think the vaccine was effective  6 = Did not think the vaccine was safe/concerned about side effects  7 = Someone else told me that the vaccine was not safe  8 = Had a bad experience with previous vaccinator/health clinic  9 = Had a bad experience or reaction with previous vaccination  10 = Fear of needles  11 = Not possible to leave other work  12 = Religious reasons  13 = Refusal of male head of household  14 = Belief it can lead to infertility  15 = Already vaccinated against diphtheria  16 = Other beliefs/traditional medicine  17 = Other (please specify): | Trying to learn about barriers to vaccination |
| *Diphtheria history* |  |  |
| Did this individual have suspected diphtheria during the recall period | 0 = No 1 = Yes  99 = DNK | Clear case definitions necessary |
| If yes, what was the outcome? | 1 = Recovered  2 = Still unwell (complications)  3 = Died |  |
| If still unwell, what are the long-term health problems? | 1 = Myocarditis/heart failure  2 = Neuritis  3 = Kidney failure  3 = Otitis media  4 = Problems breathing  5 = Malnutrition  6 = Other (specify) | Check all that apply  Will need community language for these, and to provide examples to guide the responders and data collectors, e.g. hoarseness of voice, regurgitation of fluid/food, weakness of limbs… |
| If yes, approximate date of disease onset? | (mm/yy) | Event calendar to help |
| If yes, were they vaccinated prior to disease onset? | 0 = No  1 = Yes (any doses)  99 = DNK | Asking again for extra certainty |
| If yes, did they have **direct contact** with another diphtheria patient within the household prior to developing symptoms? | 0 = No 1 = Yes  99 = DNK |  |
| If yes, what were your symptoms? | 1 = Whitish/greyish layer of the tongue  2 = Swollen neck  3 = Fever  4 = Sore throat  5 = Difficulty swallowing  6 = Difficulty breathing  7 = Drooling saliva  8 = Nasal regurgitation  9 = Nasal or other bleeding  10 = Malaise/Lethargy  11 = Loss of appetite  12 = Hoarseness  99 = None of the above | Check all that apply  Will need community language for these |
| If yes, was there a diagnosis by a health professional? | 0 = No  1 = Yes  99 = DNK |  |
| If yes, did they seek healthcare? | 0 = No 1 = Yes  99 = DNK |  |
| If yes, place health care sought? | 1 = Home  2 = Health centre  3 = Diphtheria Treatment Centre (*specify*)  4 = Private hospital *(specify)*  5 = Public hospital (*specify*)  6 = Traditional healer  7 = Patient Medicine Vendor/Pharmacy  8 = Other place *(specify)*  99 = DNK | Select all that apply, in order of visit |
| If yes, after seeking healthcare, where were they finally treated? | 1 = Health facility (specify)  2 = Home  99 = DNK | This is to distinguish between those receiving DTC-based and home-based care |
| How do you feel about the quality of treatment you received? | 1 = Strongly positive  2 = Quite positive  3 = Neutral  4 = Quite negative (specify)  5 = Strongly negative (specify) |  |
| If quite/strongly negative, please explain | Free text |  |
| If yes, who treated them? | 1 = MSF  2 = Other NGO (e.g. Alima/Save The Children) 3 = Government health facilities not supported by NGOs  99 = DNK |  |
| If yes, delay between disease onset and treatment beginning? | Numeric (days) | This refers to diphtheria treatment by one of the providers above (NGO/govt) |
| If no, main reason for not seeking care in a health structure? | 1 = No money/consultation too expensive  2 = Too sick to seek care  3 = Not sick enough to seek care 4 = Health facility too far away 5 = Went to traditional healer  6 = Did not have time/too busy to go 7 = No confidence in health system  8 = Security problem 9 = Care was refused at facility  10 = Refusal of male head of household 11 = Died suddenly before visit could be arranged  12 = Other reason *(specify)* 99 = DNK |  |
