## Supplementary material for "Community burden of diphtheria during the 2023–24 epidemic in Kano State, Nigeria: a population-based household survey": S1 Table. Sensitivity analysis restricted to cases diagnosed by a health professional

**S1 Table. Sensitivity analysis restricting the case definition to cases reported as diagnosed by a health professional.**

Results under the primary case definition (respondent-reported suspected diphtheria) and a restricted definition (cases whose diagnosis by a health professional was reported by the respondent).

| **Estimate** | **Primary case definition** | **Restricted definition** |
| --- | --- | --- |
| Cases, n | 88 | 58 |
| Attack rate, % [95% CI] | 1.08 [0.74,1.42] | 0.67 [0.44,0.91] |
| Ratio to the MSF-site rate (0.348%) [95% CI] | 3.1 [2.1,4.1] | 1.9 [1.3,2.6] |
| Case fatality ratio, % [95% CI] (deaths/cases) | 8.7 [3.9,18.5] (8/88) | 11.3 [4.3,26.6] (6/58) |
| Vaccination–diphtheria association, crude OR [95% CI]; p | 2.11 [1.23,3.64]; p=0.008 | 2.32 [1.07,5.02]; p=0.034 |

*AR, attack rate; CI, confidence interval; MSF, Médecins Sans Frontières; OR, odds ratio. The primary case definition is a respondent report of suspected diphtheria in a household member. The restricted definition keeps the cases whose diagnosis by a health professional was reported by the respondent; the diagnosis was not verified against records, so the restricted definition is not a confirmed case definition. Attack rates, case fatality ratios, and ORs are survey-weighted; case fatality ratio CIs are logit-transformed survey intervals. The MSF-site rate is the number of residents of the four Local Government Areas notified as diphtheria cases at three MSF-supported treatment sites with symptom onset from 1 January 2023 to 25 October 2024, the survey recall period (9,864; notification date for 11 cases with no onset date), divided by the population of the four Local Government Areas (2,837,111) (MSF line list; aggregated counts in S2 Table). It excludes cases notified at other facilities. The CI of the ratio reflects survey sampling uncertainty only. The positive association between reported vaccination and diphtheria remained under the restricted definition.*
