## Supplementary material for "Community burden of diphtheria during the 2023–24 epidemic in Kano State, Nigeria: a population-based household survey": S2 File. Study protocol (version 2024-07-08)

Retrospective study of diphtheria outbreak

Kano state, Nigeria

2023-2024

Survey protocol

Version 2024-07-08

Jonathan Polonsky

| **Version** | Version 2024-07-08 |
| --- | --- |
| **Survey design** | Retrospective survey with retrospective cohort study |
| **Survey period** | July-Aug 2024 (1 month) |
| **Survey site** | Kano state, Nigeria |
| **Principal investigators (Study coordinator)** | Jonathan Polonsky, Epicentre |
| **Local Partner Institutions** | Kano State Ministry of Health |
| **Co-investigators** | **Jonathan Polonsky,** Epicentre, Paris, France  **Suleiman Hudu,** MSF WaCa, Kano state, Nigeria  **Khadidja Uthman,** MSF WaCa, Kano state, Nigeria  **Philip Eghosa Evbuomwan,** MSF WaCa, Nigeria  **Hashim Juma Omer Osman**, MSF WaCa, Kano state, Nigeria  **Abdulwahab Kabir Sulaiman***,* Kano State Ministry of Health, Kano State, Nigeria  **Imam Wada Bello***,* Kano State Ministry of Health, Kano State, Nigeria  **Sulaiman Iliyasu Hamisu**, Kano State Ministry of Health, Kano State, Nigeria  **Bola Biliaminu Lawal**, Nigeria Centre for Disease Control and Prevention (NCDC), Nigeria  **Yanu Mbuyi, MSF WaCa,** Abuja, Nigeria  **Dagemlidet Tesfaye Worku,** MSF WaCa, Abidjan, Côte d’Ivoire  **Ichola Ismael Adjaho,** MSF WaCa, Abidjan, Côte d’Ivoire  **Franck Ale, Epidemiologist,** MSF WaCa, Abidjan, Côte d’Ivoire  **Etienne Gignoux,** Epicentre, Paris, France |
| **Protocol development and survey design** | Jonathan Polonsky, Epicentre, Paris, France  Suleiman Hudu, MSF WaCa, Kano state, Nigeria  Khadidja Uthman, MSF WaCa, Kano state, Nigeria  Hashim Juma Omer Osman, MSF WaCa, Kano state, Nigeria  Abdulwahab Kabir Sulaiman*,* Kano State Ministry of Health, Kano State, Nigeria  Bola Biliaminu Lawal, Nigeria Centre for Disease Control and Prevention (NCDC), Nigeria  Dagemlidet Tesfaye Worku, MSF WaCa, Abidjan, Côte d’Ivoire  Ichola Ismael Adjaho**,** MSF WaCa, Abidjan, Côte d’Ivoire  Franck Ale, MSF WaCa, Abidjan, Côte d’Ivoire  Etienne Gignoux, Epicentre, Paris, France |

### List of abbreviations

| 95% CI | 95% Confidence Interval |
| --- | --- |
| AR | Attack Rate |
| CFR | Case Fatality Ratio |
| CMR | Crude Mortality Rate |
| DSMR | Diphtheria-Specific Mortality Rate |
| DTC | Diphtheria Treatment Centre |
| GDPR | General Data Protection Regulation |
| HBC | Home-Based Care |
| IDH | Infectious Diseases Hospital |
| IPC | Infection Prevention and Control |
| IQR | Interquartile Range |
| MMSH | Murtala Muhammed Specialist Hospital |
| MSF | Médecins Sans Frontières |
| MSF-WACA | Médecins Sans Frontières – West and Central Africa |
| NCDC | Nigeria Centre for Disease Control and Prevention |
| OCA | Operational Centre Amsterdam |
| OCBA | Operational Centre Barcelona-Athens |
| OCB | Operational Centre Brussels |
| OCG | Operational Centre Geneva |
| OCP | Operational Centre Paris |
| OPD | Outpatient Department |
| RCCE | Risk Communication and Community Engagement |
| SMoH | State Ministry of Health |
| SRS | Simple Random Sampling |
| U5MR | Under 5 Mortality Rate (mortality rate in children under 5 years of age) |
| VA | Verbal Autopsy |
| WHO | World Health Organization |

### 1 Introduction

#### 1.1 Context

Nigeria, situated in West Africa, is the most populous country on the African continent with an estimated population exceeding 200 million people as of 2024. The country spans approximately 923,768 km^2^ and is bordered by Benin to the west, Niger to the north, Chad to the northeast, and Cameroon to the east. Nigeria's diverse landscape includes savannahs, tropical rainforests, and coastal plains.

Nigeria is home to over 250 ethnic groups, each with its own distinct languages, traditions, and customs. The three largest ethnic groups are the Hausa-Fulani, Yoruba, and Igbo, which collectively account for a significant portion of the population. Nigeria is linguistically diverse, with over 500 languages spoken across the country. The official language is English, inherited from British colonial rule, but various indigenous languages are widely spoken, including Hausa, Yoruba, Igbo, Fulfulde, Kanuri, and others.

Kano State, located in the northwest of Nigeria, covers an area of approximately 20,131 km^2^ and is characterized by its semi-arid climate. It is estimated to have a population exceeding 15 million people, making it one of the most densely populated states in Nigeria. The state is predominantly inhabited by the predominantly Muslim Hausa-Fulani ethnic group, with Hausa being the most widely spoken language. The state's economy is largely driven by agriculture, trade, and commerce, with Kano City serving as a major commercial hub in northern Nigeria.

*[Map of Nigeria removed from this version: the original image is third-party copyrighted material (© Nations Online Project) and cannot be published under the CC BY 4.0 licence. Kano State lies in the northwest of Nigeria.]*

Figure 1: Map of Nigeria showing national and state boundaries.

Diphtheria is a bacterial infection caused by toxigenic strains of *Corynebacterium diphtheriae*, primarily causing infection of the mucous membrane of the upper respiratory tract (1). Local bacterial growth and subsequent tissue death may create a pseudomembrane potentially leading to airway obstruction; in severe cases, disseminated toxin may cause myocarditis or peripheral neuropathy (2). The disease is typically transmitted through respiratory droplets or by direct contact with cutaneous lesions or fomites (2). The incubation period is typically 2 to 5 days, and the infectious period for respiratory diphtheria is typically 2 to 4 weeks (2,3). Diphtheria antitoxin (DAT) treatment is used to neutralize the circulating toxin in respiratory diphtheria, administered ideally 48 hours post-onset of symptoms (2,4), but due to the small risk for anaphylaxis (5), a sensitization test is frequently performed prior to administration. Antibiotic treatment is used to eliminate the bacteria, halt toxin production, and reduce transmissibility (2). Prophylactic antibiotic treatment coupled with diphtheria toxoid vaccination should be considered for close contacts (2). The inclusion of a diphtheria vaccine in childhood immunization protocols after World War II has reduced the incidence of diphtheria worldwide (6,7). However, outbreaks of diphtheria, especially large ones occurring in low-income and crisis-affected settings, are increasing, linked to disrupted health services, notably routine vaccination, and waning immunity. Recent large outbreaks have occurred among Rohingya refugees in Bangladesh, in Yemen, Venezuela, and Indonesia, among others (3,8–11).

Since 2023, there has been an ongoing, multi-country diphtheria outbreak across several countries in West Africa (Nigeria, Guinea, Niger, Cameroon, and Gabon), of which Nigeria has been the most affected, with over 27,000 suspected cases and 650 deaths reported between 1^st^ January 2023 and 25^th^ February 2024 (12). Within Nigeria, Kano state has been the most affected, with over 14,000 cases and approximately 900 deaths (Figure 2).

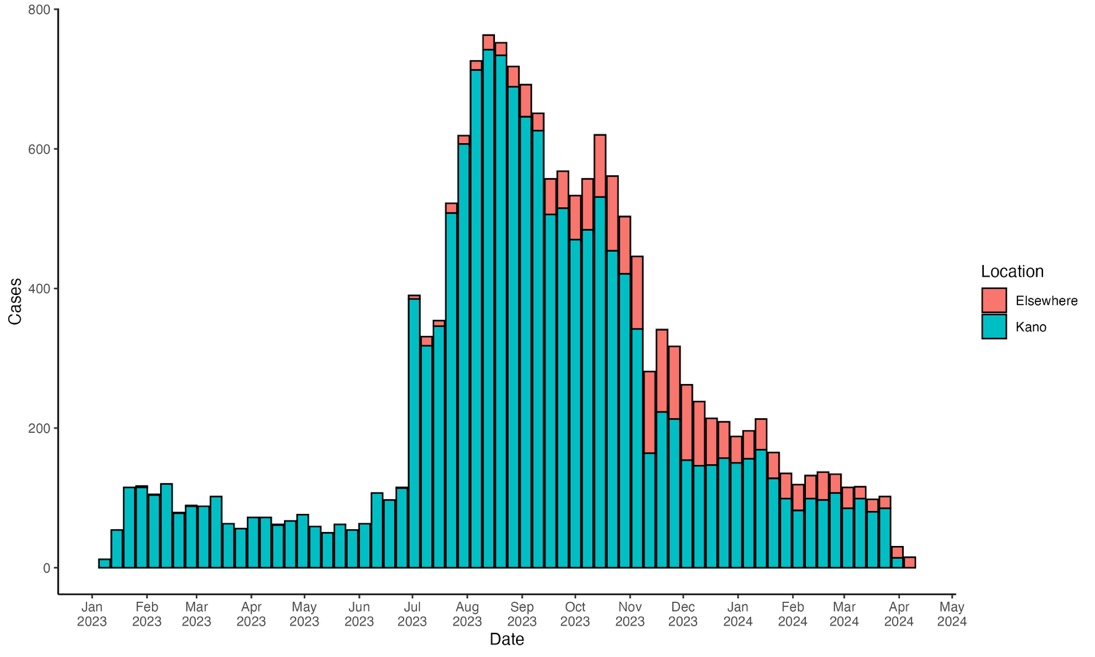

Figure 2: Nigeria national diphtheria cases (Kano state vs. all other states) reported per epidemiological week as of 10 April 2024. *Source: Médecins sans Frontières – West and Central Africa*

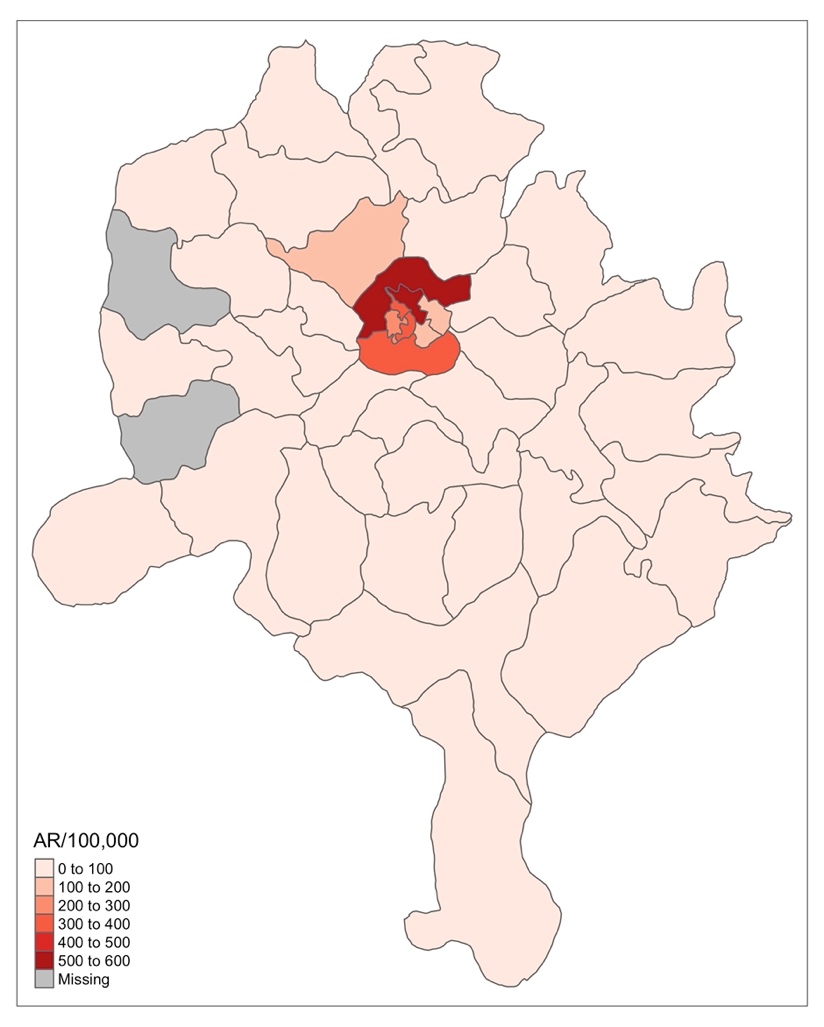

Figure 3: Geospatial distribution of diphtheria attack rate per 100,000 population, Kano state, Nigeria, 1 Jan 2023 – 10 Apr 2024. Areas in grey have missing data. Base layer: Nigeria Operational Ward Boundaries (administrative level 3), GRID3 Nigeria (eHealth Africa and Proxy Logics, 2020), <https://grid3.gov.ng/>, licensed under CC BY 4.0 (<https://creativecommons.org/licenses/by/4.0/>); boundaries were aggregated to local government area level.

In recent years, diphtheria has been re-emerging in Nigeria, due in large part to immunity gaps, particularly among children, which has resulted in high diphtheria-specific mortality rates (13–15). Nigeria has experienced several diphtheria outbreaks in recent years, the most important of which occurred between February and November 2011, when 98 cases were reported in the rural areas of Borno State, north-eastern Nigeria (16). However, the ongoing outbreak dwarfs the previous outbreaks, both in terms of caseload and mortality.

The scale of this outbreak rapidly overwhelmed the healthcare capacity to respond, and therefore Médecins sans Frontières – West and Central Africa (MSF-WACA) decided to implement a novel home-based treatment programme for the majority of less severe cases, while continuing to treat severe patients in traditional diphtheria treatment centres (DTCs). However, it is unclear to what extent this strategy was successful, whether there were any unintended consequences, and whether it would be a feasible and effective approach in future outbreaks in resource-poor settings with similar constraints. Furthermore, a better understanding of the impact of this outbreak and the factors that contributed to the high incidence and severity is critical to better respond to future epidemics in similar contexts.

#### 1.2 MSF presence in the country

Since its creation in 1973, MSF has been responding to humanitarian and public health crises in Nigeria. Currently, six operational centres (OCA, OCBA, OCB, OCG, OCP and WaCA) are delivering various health and humanitarian interventions across five of the six geopolitical zones of Nigeria. This includes emergency response to various outbreaks (including diphtheria, cholera, meningitis, COVID-19, and viral haemorrhagic fevers), and humanitarian crises precipitated by insecurity and natural disasters. In addition, MSF also delivers interventions related to maternal and child health, nutrition, Noma, and non-communicable diseases through regular projects.

MSF WaCA began operations in Nigeria in 2019, and currently has two projects in Kano and Cross River states. Since 2020, WaCA has been delivering various interventions in three thematic areas: access to primary healthcare; reproductive health; and nutrition. In January 2023, a diphtheria outbreak was declared by the Kano State Ministry of Health (SMoH), since which time MSF WaCA responded by establishing a Diphtheria Treatment Centre (DTC) at Murtala Muhammed Specialist Hospital (MMSH) in collaboration with SMoH. Due to a rapid rise in incidence, an additional DTC at Infectious Diseases Hospital (IDH) was established. In addition, a diphtheria outpatient department (OPD) and 2 contact clinics were established to provide home-based care (HBC) for patients.

During the outbreak, MSF supported all pillars of the response. These include case management/infection prevention and control (IPC); surveillance and epidemiology; vaccination (through Health Promotion campaigns); and risk communication and community engagement (RCCE). Building the capacity of the local workforce was among the key WaCA strategies to the response, and by the end of March 2024, MSF had handed over their part of the diphtheria response to the SMoH. Over the 15 months period of the intervention, a total 14,707 cases were diagnosed and manage at MSF-supported health facilities. Just over half of these (8,011, 54.5%) were hospitalized, while the remainder (6,696, 45.5%) received HBC. Among these patients, 910 were recorded as having died (CFR 6.2%).

#### 1.3 Justification for the survey

Much remains unknown about diphtheria epidemics in these contexts, including the most effective strategies to interrupt transmission and the role of health-seeking behaviour and access to healthcare. In this context, MSF has responded with a novel strategy of ‘home-based care’, treating patients at home rather than at traditional diphtheria treatment centres. It is vitally important to evaluate this approach, both in terms of effectiveness and safety, in order to assess whether this is an approach that should be implemented during future outbreaks. The intention of this study is to document the extent and impact of the outbreak, describe the long-term sequelae of disease, and evaluate the success of the home-based case strategy in terms of treatment outcome and secondary attack rates. As secondary objectives, we intend to assess the role of routine and supplementary vaccination in preventing disease, understand the barriers faced by the population in seeking and accessing quality healthcare, and determine causes of death by verbal autopsy, exploring the feasibility of conducting verbal autopsy interviews by telephone.

### 2 OBJECTIVES

#### 2.1 Overall objective

- Describe the impact of, and evaluate the MSF strategy in response to, the diphtheria epidemic in Kano state between Jan 2023 and May 2024.

#### 2.2 Specific objectives

- Estimate incidence rate, case fatality ratio (CFR), and crude- and diphtheria-specific mortality rate (CMR and DSMR, respectively) for the total population in the catchment area during the defined recall period.
- Document the long-term sequelae of disease among diphtheria survivors.
- Evaluate the performance of MSFs home-based care strategy in terms of treatment outcome and secondary attack rates within the home.

#### 2.3 Secondary objectives

- Assess vaccination coverage and estimate vaccine effectiveness in this setting.
- Estimate the coverage of the MSF diphtheria-treatment program.
- Describe typical care-seeking pathways for patients with suspected diphtheria, and identify the key barriers to seeking and accessing appropriate healthcare (both vaccination and therapeutic care).
- Determine causes of death by verbal autopsy.

### 3 Methods

#### 3.1 Study design

This study will occur in two phases using two study designs.

The first phase is a population-based retrospective survey of diphtheria incidence, mortality, and health-seeking behaviour, among a representative sample of the target population, selected using spatial sampling. A follow-up telephone-based verbal autopsy (VA) interview will be conducted to determine cause of death for all decedents identified through the household survey.

The second phase is a retrospective matched cohort study examining the effectiveness of the home-based care strategy and within-household secondary attack rates among patients treated at home and those treated as in-patients in DTCs, supplemented by review and analysis of their chart records as recorded in MSF databases.

Through a combination of these study designs, we will estimate long-term diphtheria outcomes (sequelae among survivors and delayed mortality), and healthcare-seeking behaviour and barriers to accessing care.

#### 3.2 Study area

The study will take place in Kano state, Nigeria. The household survey component of the study will take place in four LGA within Kano state, namely Ungogo, Fagge, Nassarawa, and Dawakin Tofa.

#### 3.3 Study population

The study population of the household survey will consist of all people living, or having ever lived, in the study area during the recall period. The last census was conducted in 2006 and therefore the exact population sizes are unknown, but projections estimate the 2022 Kano state population size to be 15,462,200 (17).

The study population of the retrospective cohort study will be all diphtheria patients (and their household members) residing in the study area and recorded in MSF-WACA registries during the recall period.

#### 3.4 Definitions

##### 3.4.1 Household

A household is defined as a group of people who are under the responsibility of one person or head of household and slept under the same roof the previous night.

##### 3.4.2 Head of household

The head of household is defined as follows:

- Adult household member (18 years or older) who identifies him/herself as head of household and is identified as such by the other members*, and*
- Can give accurate information on all demographic and mortality issues in his/her household (can describe with reasonable accuracy the events that occurred during the recall period), or can authorise some other household member to provide the information, *and*
- Has lived in the household the entire recall period, *and*
- Is present at the time of the survey.

A household will be excluded from the survey if none of the household members fulfil all these criteria.

##### 3.4.3 Household member

A member of the household is defined as a person who is or was part of the household according to the household definition during the recall period. This includes members of the household who died or moved out but were members of the household at any time during the recall period.

##### 3.4.4 Diphtheria patient

In the household survey, a diphtheria patient is defined as anyone reported as having been diagnosed with diphtheria. In the cohort study, a diphtheria patient is defined as anyone identified in MSF-WACA records as having had a clinical diagnosis of suspected diphtheria, whether that clinical suspicion was confirmed by laboratory test or not.

#### 3.5 Recall period

The recall period begins on January 1^st^, 2023 (New Year’s Day, a national holiday in Nigeria), and ends on the survey interview date of each individual, corresponding to at least 495 days (assuming a 01 July 2024 survey start date).

An events calendar will be developed for the chosen recall period to determine the dates of events (births, deaths, movements within the household, and care-seeking pathway) more precisely.

#### 3.6 Inclusion and exclusion criteria

##### 3.6.1.1 Household survey

A person will be included in the survey if s/he satisfies all the following criteria:

- Is or was a member of the selected household during the recall period, *and agrees to participate*
- Written informed consent has been given by the head of the household (see chapter 4.4 for details on the informed consent process).

A person will be excluded from the survey if s/he satisfies one of the following criteria:

- Refusal to participate in the survey, *or*
- Inability to locate the participant after two attempts to trace him/her, if no one in the household was entitled to answer on his/her behalf.

##### 3.6.1.2 Cohort study

A person will be included in the study if s/he satisfies all the following criteria:

- Was recorded as having been a ‘mild’ patient in the MSF database, within the catchment areas of IDH and MMSH.
- Written informed consent has been given by themselves or an appropriate respondent (such as their head of the household or primary care-giver) (see chapter 4.4 for details on the informed consent process).

A person will be excluded from the survey if s/he satisfies one of the following criteria:

- Refusal to participate in the survey, *or*
- Inability to locate the participant after two attempts to trace him/her.

#### 3.7 Sampling

##### 3.7.1 Sample size

##### 3.7.1.1 Household survey

The sample size for the survey was calculated using the following formula, implemented in R:

$$Sample size (in persondays)=\frac{\frac{expected number of deaths per day}{10000}}{\left( \frac{precision}{\frac{10000}{z}} \right)^{2}}*\left( 1+nonresponse \right)*DEFF$$

The criteria listed below were taken into consideration for this calculation of the sample size.

Table 1. Parameters for the calculation of the household survey sample size

| **Parameter** | **Value** |
| --- | --- |
| Expected CMR per 10,000 person-days | 0.25 |
| Absolute precision (per 10,000 person-days) | 0.08 |
| Recall period (days) | 550 |
| Alpha error (z) | 0.05 (1.96) |
| Power ($\beta$) | 80% |
| Design effect | 1.5 |
| Estimated household size | 5 |
| Non-response (%) | 10 |
| Total population to be sampled | 4494 |
| Total households to be sampled | 899 |

For this survey, the main objective is to estimate the CMR. Based on an expected CMR of 0.25 per 10,000 person-days, a precision around the estimate of ± 0.08 with 95% confidence interval (95% CI; alpha error = 0.05), power of 80%, 550 days for the recall period, an average of 5 household members, and an expected 10% of non-respondents, a minimum of 899 households are to be sampled.

##### 3.7.1.2 Retrospective cohort study

The sample size for the survey was calculated using the following formula, implemented in R:

$$Sample size (in each group)=\frac{p_{1}\left( 1-p_{1} \right)+p_{2}\left( 1-p_{2} \right)}{{{(p}_{1}-p_{2})}^{2}}*{(Z_{\frac{\alpha}{2}}*Z_{\beta})}^{2}*\left( 1+nonresponse \right)$$

The criteria listed below were taken into consideration for this calculation of the sample size.

Table 2. Parameters for the calculation of the retrospective cohort study sample size

| **Parameter** | **Value** |
| --- | --- |
| Expected CFR under hospital treatment (p_1_) | 0.025 |
| Expected CFR under home-based care (p_2_) | 0.05 |
| Alpha error (z) | 0.05 (1.96) |
| Power ( $\beta$) | 80% |
| Non-response (%) | 10 |
| Sample size in each group | 496 |
| Total sample size | 992 |

For this study, the main objective is to compare the proportions of patients who had severe outcomes (death or long-term sequelae) between those treated at DTCs and those treated under home-based care. Based on an expected CFR under hospital treatment of 2.5%, expected CFR under home-based care of 5%, alpha error of 0.05, power of 80%, and an expected 10% of non-respondents, a minimum of 496 patients are to be included in each exposure group.

##### 3.7.2 Sampling procedures

##### 3.7.2.1 Household survey

A spatial sampling method will be followed (rooftop points with radius). The sampling procedures will require a preparatory step to define the area of interest with one or more geo-referenced polygons. Random coordinates will be then drawn within the polygon/s and superposed to remote sensing satellite images, using the GeoSampler application developed by Epicentre (<https://apps.msf.fr/epiGeoSampler/>). The coordinates corresponding to a roof will be kept and the households living under those roofs are those selected. Coordinates not corresponding to a roof will be discarded. The number of coordinates generated in this way will be 10% of the total number of households to be included, as a second stage will occur in which nine additional rooftops will be selected on the ground according to their proximity to the randomly generated coordinates. If large multi-story or multi-family buildings are present in the sampled rooftops, interviewers will draw the households to visit within the building using a random number generator on their tablets. This process will thereby create clusters of 10 households which is for logistical considerations (90 clusters in total). The source of satellite images will be © Airbus DS (2024) from images dated 2024. Selected households will be found with the help of the OsMand app.

##### 3.7.2.2 Retrospective cohort study

Simple random sampling (SRS) will be used to select patients for inclusion in this component. Using the complete list of patients treated by MSF, patients will first be separated into those who received treatment at DTCs and those who received home-based care. From these lists, 496 patients who had been classified as “mild” will be selected by SRS from the list of patients who received treatment at DTCs, using a randomisation procedure built into R software (18). An equal number of patients who received home-based care will be randomly selected using the following matching variables: sex, age group (within five-year age bands), and geographic location of residence (admin level 3 [Ward] if possible, otherwise admin level 2 [LGA]).

##### 3.7.2.3 Verbal autopsy

All decedents identified through either study design will be eligible for inclusion in the verbal autopsy study. As telephone coverage is good throughout Kano State, it is not envisaged that this will affect the recruitment process. However, home visits will be made in the case where respondents are unreachable after three attempts in order to account for difficulties in service coverage. The verbal autopsy component of the study will be conducted at least one month after the end of the household and cohort studies, to permit this minimum amount of time to pass following a death to avoid undue distress to recently bereaved family members.

#### 3.8 Procedures

##### 3.8.1 Introduction of the survey to the community

During the preparation, the survey coordinator will share information and discuss the study with key authorities and community leaders/ groups to ensure mutual understanding and the smooth and respectful implementation of the study. The survey coordinator will also inform the administrative authorities and community leaders/representatives of the visit of the interviewer team of the selected camps several days in advance (Appendix 3).

The purpose of the survey will be explained with the help of an information notice to the administrative authorities and verbal authorisation will be obtained before starting the interviews. The survey coordinator will clearly inform the administrative authorities and community leaders/representatives that they are freely allowed to decline the participation without any consequences or penalty.

##### 3.8.2 Introduction of the survey to the households

In each selected household, the interviewer teams will explain the purpose of the survey in the language he/she is familiar with and written consent, as required by the local ethics committee, will be obtained to conduct the interviews (the consent form is presented in Appendix 2).

If the head of the household declines to participate, the interviewer team will record this information, thank the head of the household, and proceed with the subsequent household selected.

If the head of the household provides consent to participate, the interviewer team will agree with the head of the household to identify a place where the interview will be carried out with adequate confidentiality. The head of the household may propose another household member to respond to the question on behalf of all household members. The interviewer team will encourage the head of the household to discuss with the other adult members whether they agree he/she gives consent on their behalf, and whether anyone would like to answer the questions by themselves. In any case, a parent/caregiver will respond on behalf of children. The interviewer team will then explain which information will be collected for each member of the household.

#### 3.9 Data collection

##### 3.9.1 Household survey

Interviewers will fill a Kobo Collect questionnaire form for each eligible member in selected households.

If a selected household is not be available after two visit attempts (morning and afternoon), or is not willing to respond, that household will not be replaced.

The interview will be based on a standardised questionnaire that consists of the following sections:

- **Socio-economic status**: Asset ownership to explore the association of socio-economic status with outcomes of interest.
- **Household demography**: Demographic information (age and sex) and household membership history (present throughout, arrived, born, left, and/or died) of all household members during recall period.
- **Mortality**: Date and presumed cause of death, and health-seeking behaviour prior to death, for all members of the household who died during the recall period.
- **Vaccination**: Primary caregiver attitudes towards vaccination, and diphtheria vaccination status and timing for all household members.
- **Diphtheria history**: Whether each household member had suspected diphtheria during the recall period, and if so:
  - Approximate timing of disease onset.
  - Vaccination status and timing (questions asked again, for added certainty).
  - Health-seeking behaviour and treatment pathway.
  - Symptomatology.
  - Outcomes (including long-term sequelae).

A questionnaire template is provided in Appendix 7.

A specific training of the interviewer teams will be carried out to allow the interviewers to familiarise themselves with the questions, the way to ask the questions, and the mode to record them into the electronic device questionnaire form.

No personal identified data (name, telephone number, date of birth) will be collected during the survey, reducing the risk that participants will be identifiable after the survey has been completed. The address and/or the geographical coordinates that were used to sample the household will be kept and managed separately from the survey questionnaire (more details are given in chapter 4.6).

##### 3.9.2 Retrospective cohort study

Interviewers will fill a Kobo Collect questionnaire form for each selected diphtheria patient (or their care-giver).

If a selected respondent is not be available after two visit attempts (morning and afternoon), or is not willing to respond, they will be excluded from the survey.

The interview will be based on a standardised questionnaire that consists of the following sections:

- Household composition.
- Diphtheria vaccination and disease status and timing of all household members.
- For all household cases:
  - Symptomatology.
  - Outcomes (including long-term sequelae).
  - Health-seeking behaviour.

A questionnaire template is provided in Appendix 8.

A specific training of the interviewer teams will be carried out to allow the interviewers to familiarise themselves with the questions, the way to ask the questions, and the mode to record them into the electronic device questionnaire form.

The clinical data of those patients included in this cohort study, that has been previously recorded and stored in MSF databases, will be analysed for further details and verification of their clinical presentation and treatment history, and to provide a comprehensive analysis of analysis of clinical outcomes and their association with risk and protective factors.

##### 3.9.3 Verbal autopsy

A team of female interviewers will be trained specifically for this component. Interviewers will fill a Kobo Collect questionnaire form for each decedent detected either through the household survey or the retrospective cohort study. The respondent will be the decedent’s primary care-giver.

If a selected respondent is not be available after three telephone attempts, an attempt will be made to visit their household to conduct the verbal autopsy. If that selected respondent is still not available after two attempts to visit their household, or is not willing to respond, they will be excluded from the survey.

The verbal autopsy interview will be conducted using the standardized 2016 WHO instrument (19) to understand the circumstances of the death and determine the most probable cause of death. There are three questionnaires: for adults, children (1 month to 11 years) and neonates (less than 4 weeks).

##### 3.9.4 Data management

Data will be digitally recorded directly into a mobile data collection device. A dedicated electronic template on Kobo Collect will be prepared and tested during the pilot phase of the survey. Data stored in the electronic device are encrypted and transferred directly to a dedicated server located at MSF-OCP in Paris, France. The server in place is already set to protect the electronic records, and provision for technical support during the survey will be ensured. Once the survey has completed, the electronic database will be stored at Epicentre level for 5 years and then permanently deleted.

Access to the electronic database and paper consent forms will be restricted to the co-investigators of the survey and the Medical Coordinator. Other local research partners may have access after the approval of the principal and partner institutions and with the appropriate legal agreements.

#### 3.10 Data analysis

Before any analysis, data will be checked for inconsistencies in data entry and responses.

Data analysis will be conducted using R (18).

The cluster design will be taken into account by using the *survey* package with the mini-cluster variable as the primary sampling unit.

Categorical variable indicators (e.g. sex, symptomatology, cause of death) will be presented as proportions with 95% confidence intervals (95%CI). Continuous variables (e.g. age, treatment delays) will be presented as means with associated standard deviations, if normally distributed, or as medians with associated 25th and 75th interquartile ranges (IQR), if not normally distributed. Some continuous variables may be summarised in categories (e.g. age-groups) and presented accordingly.

Mortality rates during the defined recall period will be calculated using the following general formula:

$$Mortality rate=\frac{total number of deaths during the defined period}{total person\_time during the defined period}$$

For DSMR, the number of deaths due to diphtheria and person-times of the entire population will be used. For crude mortality rate (CMR), the total number of deaths and person-times of the entire population will be used. For Under-5 mortality rate (U5MR), only deaths and person-times of this specific age-group will be used in the formula. Ninety-five percent confidence intervals will be calculated and adjusted for the design effect.

The person-times will be expressed as number of deaths per 10,000 person-days and will be calculated by summing the exact number of time units that each household member passed during the recall period. For the members who were present at the beginning and are still present at the end of the recall period the total time units of the recall period will be accounted. For the members who were not present either at the beginning or at the end of the recall period (i.e., new-borns, new arrivals, deaths and departures) a fine calculation of the time units passed in the household will be counted (for example the person-times of a new-born will be the number of days computed as the date of the survey minus the date of birth).

Estimates of actual design effect will also be calculated for each variable and those with effects greater than 1 will be reported. Where appropriate, differences in proportions will be tested using the Pearson χ2 test, differences in means will be tested using the Student’s t-test if normally distributed or the Wilcoxon rank-sum test if not normally distributed. Associated p-values (p) will be also presented.

### 4 Ethical considerations

#### 4.1 Ethical references

The survey will be conducted in accordance with the Declaration of Helsinki (20), the International Ethical Guidelines for Epidemiological Studies (21) and the International Ethical Guidelines for Health-related Research Involving Humans (22). The Principal Investigator is responsible for ethical compliance of the survey.

#### 4.2 Ethical approval

The MSF Ethics Review Board approved the standardised survey protocol used as the basis for this study protocol. This protocol will be submitted to the MSF Ethics Review Board and the Kano State Ministry of Health Ethics Committee for approval.

#### 4.3 Administrative endorsement

The ethics approved copy of this protocol, together with an overall explanation of how the survey will be carried out, will be presented to the local administrative authorities and the survey will be carried out after their written endorsement.

The community key persons in the survey area such as camp heads, religious leaders, and opinion makers, will be informed about the purpose of the survey and their agreement will be sought i.e., by organising a community meeting to present the survey.

#### 4.4 Written informed consent

#### 4.4.1 Household survey

Written informed consent (Appendix 2), as required by the local ethics committee, will be sought from every head of the selected households after providing a written information notice (Appendix 1) translated into a local language (Hausa) and oral information about the study objectives, study procedures, and a clear explanation of the risks and benefits derived from participation in the research.

The head of the household may choose to delegate answering the questionnaire to another member of the household who is more informed regarding the questions that will be asked.

All participants included in the surveys will have the investigations explained to them in a language (Hausa) with which they are familiar. All participants will be informed explicitly that they have the right to withdraw from the study at any time during the research or to decline to answer to some questions without any negative consequences and no incentives or inducements will be provided to any respondents. It will be made clear that everyone approached for the survey is completely free to participate or not.

A copy of the information notice should always remain within the household after conducting the interviews. Copies of the Information notice, and the signed informed consent in the local language (Hausa) will be given to the participant. The originals will be filed in a locked filing cabinet. Consent will be kept for five years after the completion of the research, in a secured and confidential, locked archive at the local MSF office.

For illiterate participants, a literate witness who is not part of the study team and chosen by the participant will be present throughout the information process and co-sign the informed consent form to confirm that the participant has understood the provided information and agrees voluntarily to participate in the study. Provision of written informed consent will be done in an area where sufficient privacy is ensured. Illiterate participants will express their consent by thumbprint.

#### 4.4.2 Retrospective cohort study

Written informed consent, as required by the local ethics committee, will be sought from every participant after reading an information notice (Appendix 3) translated into a local language (Hausa) and including information about the study objectives, study procedures, and a clear explanation of the risks and benefits derived from participation in the research.

All participants included in the study will have the investigations explained to them in a language (Hausa) with which they are familiar. All participants will be informed explicitly that they have the right to withdraw from the study at any time during the research or to decline to answer to some questions without any negative consequences and no incentives or inducements will be provided to any respondents. It will be made clear that everyone approached for the study is completely free to participate or not.

#### 4.5 Risks and benefits for participants and contingency plans

The retrospective mortality survey does not cause any physical harm to participants. Nevertheless, asking for details of recent deaths in the household may be upsetting and/or intrusive, particularly in this setting. The interviewers will receive a training on first psychological aid by a member of the medical team, including referral paths for severe cases.

In addition, community members may believe that mortality questions are asked as part of investigation the involved parties and thus endanger both the participants and the research team. Mitigation measures for this will include briefing the interviewers on the MSF principles of independence, neutrality, and impartiality and this will be explained clearly in the information sheets (for community leaders and heads of households), and during meetings with community leaders and emphasized during the interviews that the aim of these questions is to document the diphtheria burden on these communities.

There is also the risk to communities of breach of confidentiality and/or stigmatisation at community level.

There will be no direct benefit for the participants. However, over the course of the survey, emergency cases identified during interviews will be referred to the nearest healthcare facility, with free care supported either by MSF or another organization.

In addition, the community may benefit from the survey results:

- A better understanding of the rates and causes of death in the area will facilitate more appropriate program development and a more efficient use of resources.
- Specific data regarding mortality, violence, and malnutrition and estimates of the causes of mortality are crucial for appeals at the national and international levels.
- A better understanding of the main barriers to health-seeking behaviour related to diphtheria treatment.
- The data collected will enable a better understanding of the results of the MSF interventions, in particular the home-based care strategy, evidence for which is currently lacking.

#### 4.6 Confidentiality

Participants’ privacy will be respected during the interview process. Staff will be trained in how to determine appropriate conditions for maintaining confidentiality during the interview process, including choosing the best location when the context makes it difficult to have privacy (for example, a single room dwelling).

The confidentiality of the data collected from the participants will be ensured both during and after the survey. Participant names will not be recorded on questionnaire, and individual person records will be linked only to a household number throughout the data entry and analysis process. Any data that could be combined with other data sources to make individual records potentially identifiable will not be distributed outside the survey location or appear in any report or publication.

All geographical coordinates will be used for sampling purposes only. They will then be deleted once the data have been collected, the database created, and the analysis completed.

An electronic database will be generated from the electronic questionnaires created in the Kobo Collect application. Neither the information collection materials nor the electronic database will contain names, addresses, or other personal information about participants to ensure their anonymity. Data will be de-identified when the database is created, and questionnaires will be deleted once the database is created. This password-protected electronic database is compliant with the General Data Protection Regulation (GDPR) and will be stored on a secure server at MSF-OCP in France for 5 years after the survey is completed and then permanently deleted.

### 5 Collaboration

This survey will be carried out in collaboration between Epicentre, MSF-WACA, Kano SMoH, and the Nigeria Centre for Disease Control and Prevention (NCDC). All collaborating partners have provided substantive inputs to the survey protocol, and will participate in the training of surveyors and contribute to any scientific publications that result from this study. The collaborating partners will try their best to involve the community in the preparation and implementation of the survey and the dissemination of the results.

Epicentre is the implementing agent and oversees the entire survey in collaboration with MSF. Epicentre is also in charge of the field activity supervision, the data analysis and of the report writing. MSF-WACA is the survey sponsor and is responsible for the funding. Permission for publication must be obtained from MSF-WACA and the MoH.

Survey results will belong to MSF-WACA and the MoH in Nigeria, the country where the survey will be conducted.

### 6 Dissemination of results

MSF-WACA commits to sharing survey results with the community surveyed. The local community will be involved and informed through meetings with the community leaders. The MSF medical team will decide about the best venues to display the results.

The MSF teams project, coordination and headquarters will be given the final report and meetings will be held with humanitarian affairs and communications teams to discuss the best way for them to use the indicators for their activities (e.g., press release in national and/or international media). The MoH and other health and humanitarian organisations working in the region will also be given a copy of the report. Scientific articles will be also considered as a means of wider dissemination.

### 7 Field implementation

#### 7.1 Selection and tasks of the interviewer teams

Dedicated teams of two interviewers each will collect the necessary data for the survey. We estimate that 12 teams will be able to complete the data collection in 20 days (see chapter 7.6).

Job application advertisement and distribution will be done under the national work regulations. Interviewers will be selected according to the following criteria:

- Able to read and write in English, and
- Fluent in the local languages (Hausa), and
- Available for the entire time of the survey (training and interview days), and
- Motivated to participate in the survey, and
- Have no conflict of interest, and
- Experience with interviews in difficult settings and survey populations would be an advantage, and
- Expected to be accepted by the population

#### 7.2 Supervision

The principal investigator is the overall responsible for the final version of the protocol, overall quality of the survey and data analysis, and the final report.

The principal investigator will ensure that the following tasks are performed:

- Preparation of all necessary documents (protocol, electronic questionnaire, informed consent forms).
- Secure the necessary ethics approvals (the exemption from the MSF ERB review from the MSF-WACA Medical Director and the approval of the Kano State Ministry of Health Ethics Committee in Nigeria.
- Preparation of the field component of the survey (training of the survey teams, logistics, materials) together with the MSF team in the field.
- Follow-up of the field component of the survey.
- Validation of data entry.
- Data analysis.
- Report writing.
- Ensuring ethical compliance during implementation of the survey through supervision and training.

#### 7.3 MSF field support

- Administrative support for survey preparation at the field level and during field part, such as presentation of the survey protocol to the relevant national or SMoH ethics committee or institutional review board, payment of survey teams.
- Human resources support, such as hiring survey team/interviewers.
- Logistic support for survey preparation at the field level and during field part, such as organizing sufficient cars including drivers for the field part of the survey, providing communication tools and MSF ID (e. g. aprons, vests, or arm bands) to the survey teams, stationary, printing the questionnaire and consent forms.

#### 7.4 Training and pilot phase

A three-day training will be given to all interviewers to familiarise them with the background of the survey, the questionnaire, the information sheet, and the informed consent procedures. The training will be given in English, with translation if needed, by the principal investigator. It consists of an intensive review of the questionnaire and the information sheet including role-plays. Additional training will be provided on important ethical considerations regarding the protection and respect of the autonomy and confidentiality of participants and their responses. Interviewers will also be trained to identify those health conditions that indicate medical referral to the nearest appropriate health facility, and the correct process for making such a referral.

As the interviews will be held in the national language, the principal investigator should ensure that all interviewers are using the same and correct wording for providing information to the households and for the interviews.

The questionnaire will be tested in the field, in households in an area not selected for the survey, during a pilot day, and will be improved if needed. The selected area will be made aware of the survey the day before the teams arrive. Approximately 15 households will be tested by each team. Written consent for participation in the pilot will be sought in the same way as for the survey. The data collected during the pilot phase will be used solely for identifying issues in the process and will not be included in the survey data. These data will be destroyed within seven days of collection.

A preparation meeting will be held with associated staff such as logisticians, community liaisons and drivers to explain the overall survey and their roles and expectations.

During the training, investigators will ensure that the interviewers go through the following points:

- Identification of the sampled households according to the sampling method, including use of OsmAnd, and the protocol in case a household cannot be identified.
- Introduction to the household and to the head of the household (including how to properly go through the process of written consent).
- Identify a suitable location for the interview.
- Interview (including standardization regarding posing of questions, empathy, and patience for sensitive topics).
- Concluding the interview.

#### 7.5 Safety and security of interviewers

During the training, interviewers will be briefed on security, and will follow all along the field activity the MSF security rules that are implemented in the project. The investigators will check with the MSF referent person, the security status every morning, before sending the interviewers to the field. A communication system will be put in place so that interviewers will be reachable, while in the field, and can rapidly return to the MSF premises, if necessary.

#### 7.6 Timeframe in the field

A preliminary plan of the field part of the survey is presented in the table below.

Table 3. Preliminary plan of the field component of the household survey, Kano state, Nigeria, 2024

| **Activity** | **# days** | **Dates** |
| --- | --- | --- |
| Final preparation of the survey | 3 | July 2024 |
| Training and pilot study | 4 | July 2024 |
| Data collection | 20 | July 2024 |
| Buffer | 3 | July 2024 |
| **Total** | **30** | **July 2024** |

### 8 OPERATIONAL Budget

| **Description** | **Number** | **Cost per unit** | **Volume** | **Total amount** | **Total amount** |
| --- | --- | --- | --- | --- | --- |
|  |  | **Euros** |  | **Euros** | **Nigeria Naira** |
| **Human Resources** |  |  |  |  |  |
| Principal investigator (visa, plane ticket, *per diem*) | 1 |  | 2 weeks | 4,500 | 7,425,000 |
| Supervisor | 2 | 25/day | 16 days | 800 | 1,320,000 |
| Surveyors | 24 | 15/day | 16 days | 5760 | 9,504,000 |
| Verbal autopsy surveyors | 5 | 15/day | 4 days | 300 | 495,000 |
| **Operational Costs** |  |  |  |  |  |
| Document translations | 1 | 200 | - | 200 | 330,000 |
| Phone credit | 16 | 10 | - | 160 | 264,000 |
| Telephones (Kobo) | 16 | 150 | - | 2400 | 3,960,000 |
| Survey packs (backpack, water bottle, stationary, pens, etc) | 14 | 20 | - | 280 | 462,000 |
| **Transportation** |  |  |  |  |  |
| Rental cars with fuel for survey | 6 | 50/day | 16 days | 4800 | 7,920,000 |
| **Training (remainder)** |  |  |  |  |  |
| Training cost (per diem, room rental) | 1 | 300/day | 3 days | 900 | 1,485,000 |
| Water bottles for survey team, backpack etc | 1 | 10 /day | 16 days | 160 | 264,000 |
| Dissemination of results | 1 | 3000 | 1 | 3000 | 4,950,000 |
| **TOTAL** |  |  |  | **23,260** | **38,379,000** |

### 10 Appendixes

Appendix 1: Household survey information sheet (Participant)

**Information Sheet**

**Survey title:** Retrospective study of diphtheria outbreak, Kano state, Nigeria, 2023-2024

**Study sponsor:** Médecins Sans Frontières

**Study site:** Ungogo, Fagge, Nassarawa, and Dawakin Tofa LGAs, Kano state, Nigeria

**Coordinating Institutions:**

**Médecins Sans Frontières (**B222 Shehu Kazaure Road, Hotoro GRA, Kano, Nigeria**)**

**Epicentre (**34 Av. Jean Jaurès, 75019 Paris, France**)**

**Kano State Ministry of Health (**No. 4 Post Office Road, Kano, Nigeria**)**

**Introduction**

Dear Sir/Madam,

Thank you for taking the time to listen to our information about this survey.

My name is ............... and I work for Médecins Sans Frontières, known also as MSF.

As you might already know, MSF jointly with the Ministry of Health is providing health care in Kano state.

Your family was selected to participate in a survey to evaluate the health conditions of the population living in Kano state. The survey will be carried out from approximately July 1^st^ to 15^th^, 2024.

Before you decide whether to participate, it is important that you understand the contents of this information notice, which explains how we are carrying out the survey.

You can read this notice, or have it read to you, and we will explain it to you. Please ask us to stop as we go through the information if you have any questions. We will answer any questions you may have now or later.

**What is the survey about?**

With this survey we aim to knowing better the health problems in your community, so that we can improve our work as a medical organisation especially concerning the current health conditions. To do so, we will ask questions related to recent deaths that occurred in your family, including deaths that occurred because of tragic events. Secondly, MSF might use the results of this survey to advocate on behalf of the population at regional, national, or international level in order to raise awareness of the situation in your community.

Your participation to the survey will consist of answering to questions such as:

- The number of persons who has been living in household during a certain period, including member who are not there anymore, either because they left, or died.
- The relation of each household member to the head of the household.
- The age and sex of each household member.
- For those members who died since January 2023, the reason and place of death (if known).
- For those members who had suspected diphtheria since January 2023, some further details about their healthcare choices and the final outcome of their disease.
- The diphtheria vaccination status of all household members.

The whole process should take about 20-25 minutes. Data will be digitally recorded directly into a dedicated phone/tablet by the interviewer team.

**Why is my household being asked to participate?**

Ideally, we would like to speak with every household in the community, but this would require a lot of time. So, we opted to select only a few households in each village in a ‘random’ way, which means that they are selected by chance or coincidentally. And your household is one of those selected.

**What happens if I agree to let my household participate in the survey?**

If you agree, we will ask to you – or to a person that you designate – to answer to the questions we mentioned above. Note that the surname of your family nor the name of any family members will not be recorded anywhere. However, a unique survey ID, which is a number, will be assign to the household. This number will be used exclusively to carry out the data analysis.

You or another member of your family may find it distressing to respond to deaths. We ensure we will give you time, and we will stop the interview if you feel to do so. And you are free to decline to answer any questions and to withdraw or stop the interview at any time without any negative consequences to you or your family now or in the future. We will refer you or your family member to the mental health support services available in the site if you wish so. Please be aware that, if a death is reported, a follow-up interview will be conducted by telephone to better understand the circumstances of the death. At that time, you will again be provided with information regarding the study purposes and given the opportunity to agree or refuse to participate.

**Do I have to let my household participate?**

The participation in this survey is voluntary. If you do not agree to participate, we will not ask you any question and we move on with the other households that have been selected. Moreover, even if you agree, you can refuse to answer to some or all questions that will be asked or to stop the interview at any time.

**What are the risks and benefits of participation?**

As with any collection of data, there is always a risk of loss of confidentiality of personal data. However, getting asked personal questions or giving out personal information may be embarrassing or upsetting for you. You can refuse to answer certain questions without any consequences. There is also a risk related to the loss of confidentiality. To decrease this risk, we will work with local personnel who will be meticulously trained in interview techniques and study related documents will be kept in secured in a locked cabinet accessible by the study team only. Neither you nor your family will receive any direct benefit such as food or payment as a reward for participating. However, if someone is ill in your household, he/she will be referred to of our health centres which will be free.

**How will privacy and confidentiality be assured?**

Information collected will be kept confidential and used only for the purpose of the survey. This means we will not ask you for the surname your household, nor the name of your household members. GPS points will be used to identify the randomly selected households the day of the survey but will not be used otherwise. In addition, all documents (both paper and electronic) will be kept in a secure location accessible only to team members. The survey site locations may be used in anonymised analyses in the survey report. The interview will be conducted in the participant household with no interruption to ensure privacy.

After the survey is completed, all paper documents will be archived in a secure room at MSF in Kano state for 5 years and then permanently destroyed. Electronic data will be stored on a secure MSF-OCP server in the European Union. After 5 years the electronic database and all electronic survey data will be permanently deleted.

**How will the results be used?**

Once we have the results of this survey, we will organize meetings with all communities that participated in the research, to inform them of the results.

MSF might also use the results of this survey to advocate on behalf of the population at regional, national, or international level to raise awareness of the situation in your community and scientific articles and publications will be also considered as a means of wider dissemination.

Do you have any questions? Please feel free to ask us, we are happy to answer.

In case you have any questions you are free to contact the survey supervisor or the principal investigator whose contacts are indicated below:

Suleiman Hudu at [telephone number removed]

Jonathan Polonsky at [email address removed]

If you have any questions or need further information, you are free to contact the

- MSF office at No. 222 Shehu Kazaure Street, Hotoro GRA, Kano state, [telephone number removed], or
- you may also contact our Data protection officer in case you would like to withdraw your data from the study at
- Kano State Ministry of Health, Post Office Road, Kano state, [telephone number removed]

#### Appendix 2: Household survey written consent form

**Written Consent Form**

**Survey title: Retrospective study of diphtheria outbreak, Kano state, Nigeria, 2023-2024**

I __________________________________have been invited to participate in a study about the health conditions of the people living in Kano state. I have read the above information, or it has been read to me. I have had the opportunity to ask questions about the study and any questions that I have asked have been answered to my satisfaction. I have understood the objectives and purposes of this study, as well as the risks and benefits of participation in this study.

I voluntarily consent to participate in this study and I understand that I have the right to withdraw my participation in the study at any time, without giving any reason and without this affecting my or my family’s care for now and in the future.

I agree that the data recorded during the study may be processed automatically by the responsible of the study or his representative. I understand that all data and study documents will be archived for 5 years and then destroyed. I understand that during this period, I may at any time request access to my data, receive a copy, correct, or complete them. I understand that I can ask the study coordinator to delete all data.

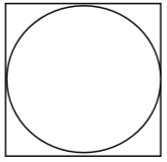
I understand that to participate in the study, I must sign or print my name below and that a copy of this information sheet and consent form will be provided to me.

**Participant signature**: _________________________

Date: d|__|__| m|__|__| y|__|__|__|__|

(or fingerprint, if illiterate)

If the head of household is illiterate

Witness declaration: I certify that the information in the information sheet and consent form was accurately explained to the participant. I confirm that the participant has had the opportunity to ask questions, and that he/she freely gave informed consent. *(This person should be chosen by the participant and must not have any connection with the research team)*

**Witness name**: _______________________**Witness signature**: _____________________

Date: d|__|__| m|__|__| y|__|__|__|__|

**Interviewer declaration:** I explained the purpose of the study to the participant and answered all his/her questions. To the best of my knowledge, he/she understands the purpose, procedures, risks and benefits of this study and voluntarily accepts to participate in the study.

**Interviewer name**: _________________________ **Interviewer signature**: _________________________

Date: d|__|__| m|__|__| y|__|__|__|__|

A copy of this information sheet and informed consent form was provided to the participant

#### Appendix 3: Household survey information sheet (Village/Neighborhood Leader)

**Information Sheet**

**Survey title:** Retrospective study of diphtheria outbreak, Kano state, Nigeria, 2023-2024

**Study sponsor:** Médecins Sans Frontières

**Study site:** Ungogo, Fagge, Nassarawa, and Dawakin Tofa LGAs, Kano state, Nigeria

**Coordinating Institutions:**

**Médecins Sans Frontières (**B222 Shehu Kazaure Road, Hotoro GRA, Kano, Nigeria**)**

**Epicentre (**34 Av. Jean Jaurès, 75019 Paris, France**)**

**Kano State Ministry of Health (**No. 4 Post Office Road, Kano, Nigeria**)**

**Introduction**

Dear Sir/Madam,

Thank you for taking the time to listen to our information about this survey.

My name is ............... and I work for Médecins Sans Frontières, known also as MSF.

As you might already know, MSF jointly with the Ministry of Health is providing health care in Kano state.

Your village was selected to participate in a survey to evaluate the health conditions of the population living in Kano state. The survey will be carried out from approximately July 1^st^ to 15^th^, 2024.

Before you allow us to pursue the survey in your village, it is important that you understand the contents of this information notice, which explains how we are carrying out the survey.

You can read this notice, or have it read to you, and we will explain it to you. Please ask us to stop as we go through the information if you have any questions. We will answer any questions you may have now or later.

**What is the survey about?**

With this survey we aim to understand better the health problems in your community, so that we can improve our work as a medical organisation especially concerning the current health conditions. To do so, we will ask questions related to recent deaths that occurred in your family, including deaths that occurred because of tragic events. Additionally, MSF might use the results of this survey to advocate on behalf of the population at regional, national, or international level in order to raise awareness of the situation in your community.

Your participation to the survey will consist of answering to questions such as:

- The number of persons who has been living in household during a certain period, including member who are not there anymore, either because they left, or died.
- The relation of each household member to the head of the household.
- The age and sex of each household member.
- For those members who died since January 2023, the reason and place of death (if known).
- For those members who had suspected diphtheria since January 2023, some further details about their healthcare choices and the final outcome of their disease.
- The diphtheria vaccination status of all household members.

The whole process should take about 20-25 minutes.

**Why is my village being selected for the survey?**

Ideally, we would like to speak with every household in every community in Kano state, but this would require a lot of time. So, we opted to select only a few households in each village in a ‘random’ way, which means that they are selected by chance or coincidentally. Your village is one of those selected.

**What happens if I agree for the survey to take place in my village?**

If you agree, we will select randomly about 16 households in your village and seek for consent from each head of household. If they agree, we will ask questions about the current diphtheria outbreak and its burden on the household. Note that we do not collect any name or surname. Each interview will be conducted by trained staff and should not last more than 20 minutes.

**Does my village have to participate?**

The participation of your village in this survey is entirely your decision. You are free to accept or not. If you do not want your village to participate, this will not affect the usual medical care that you or your village will receive in the health centres, now or in the future.

If you agree, there will be no compensation for your participation in this survey. We will not distribute anything.

**If I agree, do the households have to participate?**

The participation of the household is voluntary, and they are free to participate in the study or not, even if you agree. If they do not wish to participate, this will not have any impact on the normal medical care that they or their families will receive in the health centres, now or in the future. If they choose to participate, they will not receive any compensation, as we will not be giving anything out.

**What are the risks and benefits of participation?**

This study does not pose any physical risk to the participants. Nevertheless, asking about personal information may feel intrusive and in village contexts there may be limited privacy. The interviewers will be trained to ensure the confidentiality of the discussion. To decrease this risk, we will work with local personnel and they will be meticulously trained in interview techniques. Participants may refuse to answer any questions without any consequences. The data we collect will not contain the names of the participants.

There will be no direct benefit to your village, however, by participating you will help us better understand the burden of diphtheria on the population and this will therefore allow us to put into place more appropriate healthcare programs.

The information collected will also enable us to better understand the diphtheria epidemic, as well as the actions that MSF has taken in Kano state.

Members of selected households can receive mental health support if needed. Members of selected households who are unwell at the time of visit will be referred to the nearest health facility.

**How will confidentiality be assured?**

Information collected will be kept confidential and used only for the purpose of the survey. This means we will not ask any surname nor name of any household members. GPS points will be used to identify the randomly selected households the day of the survey but will not be used otherwise. In addition, all documents (both paper and electronic) will be kept in a secure location accessible only to team members. The survey site locations may be used in anonymised analyses in the survey report.

After the survey is completed, all paper documents will be archived in a secure room at MSF in Kano state for 5 years and then destroyed. Electronic data will be stored on a secure MSF server in the European Union. After 5 years the survey ID used as identifier will be deleted from the electronic database.

**How will the results be used?**

When the study is over, the information collected will be analyzed, and community meetings will be organized to explain the main conclusion from the study in the villages that participated in the study.

MSF might also use the results of this survey to advocate on behalf of the population at regional, national, or international level to raise awareness of the situation in your community.

Do you have any questions? Please feel free to ask us, we are happy to answer.

In case you have any questions you are free to contact the survey supervisor or the principal investigator whose contacts are indicated below:

Suleiman Hudu at [telephone number removed]

Jonathan Polonsky at [email address removed]

If you have any questions or need further information, you are free to contact the

- MSF office at No. 222 Shehu Kazaure Street, Hotoro GRA, Kano state, [telephone number removed], or
- you may also contact our Data protection officer in case you would like to withdraw your data from the study at
- Kano State Ministry of Health, Post Office Road, Kano state, [telephone number removed]

The community that I __________________________________ represent has been invited to participate in a study about the health conditions of the people living in Kano state. I have read the above information, or it has been read to me. I have had the opportunity to ask questions about the study and any questions that I have asked have been answered to my satisfaction. I have understood the objectives and purposes of this study, as well as the risks and benefits of participation in this study.

I voluntarily consent for my community to participate in this study and I understand that I have the right to withdraw my participation in the study at any time, without giving any reason and without this affecting my community’s care for now and in the future.

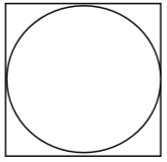
I understand that for my community to participate in the study, I must sign or print my name below and that a copy of this information sheet and consent form will be provided to me.

**Community leader’s signature**: _________________________

Date: d|__|__|	m|__|__|	y|__|__|__|__|

(or fingerprint, if illiterate)

**Interviewer declaration:** I explained the purpose of the study to the community leader and answered all his/her questions. To the best of my knowledge, he/she understands the purpose, procedures, risks and benefits of this study and voluntarily accepts for the community to participate in the study.

**Interviewer name**: _________________________ **Interviewer signature**: _________________________

Date: d|__|__| m|__|__| y|__|__|__|__|

#### Appendix 4: Retrospective cohort survey information sheet (Participant)

**Information Sheet**

**Survey title:** Retrospective study of diphtheria outbreak, Kano state, Nigeria, 2023-2024

**Study sponsor:** Médecins Sans Frontières

**Study site:** Kano state, Nigeria

**Coordinating Institutions:**

**Médecins Sans Frontières** B222 Shehu Kazaure Road, Hotoro GRA, Kano, Nigeria

**Epicentre (**34 Av. Jean Jaurès, 75019 Paris, France**)**

**Kano State Ministry of Health (**No. 4 Post Office Road, Kano, Nigeria**)**

**Introduction**

Dear Sir/Madam,

Thank you for taking the time to listen to our information about this survey.

My name is ............... and I work for Médecins Sans Frontières, known also as MSF.

As you might already know, MSF jointly with the Ministry of Health is providing health care in Kano state.

As you (or a member of your household) were a recent patient of MSF, you have been selected to participate in a study to evaluate the service provided during the illness.

Before you decide whether to participate, it is important that you understand the contents of this information notice, which explains how we are carrying out the survey. Please ask me to stop as I go through the information if you have any questions. I will answer any questions you may have now or later.

**What is the survey about?**

With this survey we aim to understand better the best way of treating diphtheria in your community and communities like yours, so that we can improve our work as a medical organisation. To do so, we will ask questions related to a recent episode of ill health. Additionally, MSF might use the results of this study to advocate on behalf of the population at regional, national, or international level in order to raise awareness of the situation in your community.

Your participation to the survey will consist of answering to questions such as:

- The outcomes of treatment.
- Your decisions regarding seeking treatment.
- Whether any other household members were also diphtheria cases.
- The vaccination status of members of your household.

The whole process should take about 20-25 minutes. Data will be digitally recorded directly into a dedicated phone/tablet by the interviewer team.

**Why am I being asked to participate?**

Ideally, we would like to speak with every person who became ill with diphtheria in the community, but this would require a lot of time. So, we opted to select only a few individuals in a ‘random’ way, which means that they are selected by chance or coincidentally. In this process, you have been selected.

**What happens if I agree to participate in the survey?**

If you agree, we will ask to you – or to a person that you designate – to answer to the questions we mentioned above.

You or another member of your family may find it distressing to discuss deaths that may have occurred. We ensure we will give you time, and we will stop the interview if you feel to do so. And you are free to decline to answer any questions and to withdraw or stop the interview at any time without any negative consequences to you or your family now or in the future. We will refer you or your family member to the mental health support services available in the site if you wish so. Please be aware that, if a death is reported, a follow-up interview will be conducted by telephone to better understand the circumstances of the death. At that time, you will again be provided with information regarding the study purposes and given the opportunity to agree or refuse to participate.

**Do I have to participate?**

The participation in this study is voluntary. If you do not agree to participate, we will not ask you any more questions. Moreover, even if you agree, you can refuse to answer to some or all questions that will be asked or to stop the interview at any time.

**What are the risks and benefits of participation?**

As with any collection of data, there is always a risk of loss of confidentiality of personal data. However, getting asked personal questions or giving out personal information may be embarrassing or upsetting for you. You can refuse to answer certain questions without any consequences. There is also a risk related to the loss of confidentiality. To decrease this risk, we will work with local personnel who will be meticulously trained in interview techniques and study related documents will be kept in secured in a locked cabinet accessible by the study team only.

Neither you nor your family will receive any direct benefit such as food or payment as a reward for participating.

**How will privacy and confidentiality be assured?**

Information collected will be kept confidential and used only for the purpose of the survey. All documents (both paper and electronic) will be kept in a secure location accessible only to team members.

After the survey is completed, all paper documents will be archived in a secure room at MSF in Kano state for 5 years and then permanently destroyed. Electronic data will be stored on a secure MSF-OCP server in the European Union. After 5 years the electronic database and all electronic survey data will be permanently deleted.

**How will the results be used?**

Once we have the results of this survey, we will organize meetings with all communities that participated in the research, to inform them of the results.

MSF might also use the results of this survey to advocate on behalf of the population at regional, national, or international level to raise awareness of the situation in your community and scientific articles and publications will be also considered as a means of wider dissemination.

Do you have any questions? Please feel free to ask us, we are happy to answer.

In case you have any questions you are free to contact the survey supervisor or the principal investigator whose contacts are indicated below:

Suleiman Hudu at [telephone number removed]

Jonathan Polonsky at [email address removed]

If you have any questions or need further information, you are free to contact the

- MSF office at No. 222 Shehu Kazaure Street, Hotoro GRA, Kano state, [telephone number removed], or
- you may also contact our Data protection officer in case you would like to withdraw your data from the study at
- Kano State Ministry of Health, Post Office Road, Kano state, [telephone number removed]

#### Appendix 5: Retrospective cohort survey written consent form

**Written Consent Form**

**Survey title: Retrospective study of diphtheria outbreak, Kano state, Nigeria, 2023-2024**

I __________________________________have been invited to participate in a study about the experiences of diphtheria treatment of the people living in Kano state. I have read the above information, or it has been read to me. I have had the opportunity to ask questions about the study and any questions that I have asked have been answered to my satisfaction. I have understood the objectives and purposes of this study, as well as the risks and benefits of participation in this study.

I voluntarily consent to participate in this study and I understand that I have the right to withdraw my participation in the study at any time, without giving any reason and without this affecting my or my family’s care for now and in the future.

I agree that the data recorded during the study may be processed automatically by the responsible of the study or his representative. I understand that all data and study documents will be archived for 5 years and then destroyed. I understand that during this period, I may at any time request access to my data, receive a copy, correct, or complete them. I understand that I can ask the study coordinator to delete all data.

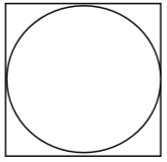
I understand that to participate in the study, I must sign or print my name below and that a copy of this information sheet and consent form will be provided to me.

**Participant signature**: _________________________

Date: d|__|__| m|__|__| y|__|__|__|__|

(or fingerprint, if illiterate)

If the head of household is illiterate

Witness declaration: I certify that the information in the information sheet and consent form was accurately explained to the participant. I confirm that the participant has had the opportunity to ask questions, and that he/she freely gave informed consent. *(This person should be chosen by the participant and must not have any connection with the research team)*

**Witness name**: _______________________**Witness signature**: _____________________

Date: d|__|__| m|__|__| y|__|__|__|__|

**Interviewer declaration:** I explained the purpose of the study to the participant and answered all his/her questions. To the best of my knowledge, he/she understands the purpose, procedures, risks and benefits of this study and voluntarily accepts to participate in the study.

**Interviewer name**: _________________________ **Interviewer signature**: _________________________

Date: d|__|__| m|__|__| y|__|__|__|__|

A copy of this information sheet and informed consent form was provided to the participant

#### Appendix 6: Verbal autopsy information sheet (Participant)

**Information Sheet**

**Survey title:** Retrospective study of diphtheria outbreak, Kano state, Nigeria, 2023-2024

**Study sponsor:** Médecins Sans Frontières

**Study site:** Kano state, Nigeria

**Coordinating Institutions:**

**Médecins Sans Frontières (**B222 Shehu Kazaure Road, Hotoro GRA, Kano, Nigeria**)**

**Epicentre (**34 Av. Jean Jaurès, 75019 Paris, France**)**

**Kano State Ministry of Health (**No. 4 Post Office Road, Kano, Nigeria**)**

**Introduction**

Dear Sir/Madam,

Thank you for taking the time to listen to our information about this survey.

My name is ............... and I work for Médecins Sans Frontières, known also as MSF.

As you might already know, MSF jointly with the Ministry of Health is providing health care in Kano state.

As you may recall, your family was previously selected to participate in a survey to evaluate the health conditions of the population living in Kano state, during which time a death in the household was reported to have occurred since 1^st^ January 2023. We are telephoning you today to ask you some follow up questions regarding the circumstances of the death, in order to better understand the risks to people’s health that occur in your community.

Before you decide whether to participate, it is important that you understand the contents of this information notice, which explains how we are carrying out the survey.

You can read this notice, or have it read to you, and we will explain it to you. Please ask us to stop as we go through the information if you have any questions. We will answer any questions you may have now or later.

**What is the survey about?**

With this survey we aim to knowing better the health problems in your community, so that we can improve our work as a medical organisation especially concerning the current health conditions. To do so, we will ask questions related to recent deaths that occurred in your family, including deaths that occurred because of tragic events. Additionally, MSF might use the results of this survey to advocate on behalf of the population at regional, national or international level in order to raise awareness of the situation in your community.

Your participation to the survey will consist of answering questions related to the death of any members of your household since 1^st^ January 2023. The whole process should take about 20-25 minutes.

**Why is my household being asked to participate?**

When your household was previously visited during a survey by MSF, a death was reported to have occurred since 1^st^ January 2023. We are asking these follow-up questions to all households that reported a death in this time period.

**What happens if I agree to let my household participate in the survey?**

If you agree, we will ask to you – or to a person that you designate – to answer to the questions we mentioned above. Note that the surname of your family nor the name of any family members will not be recorded anywhere. However, a unique survey ID, which is a number, will be assign to the household. This number will be used exclusively to carry out the data analysis.

You or another member of your family may find it distressing to respond to deaths. We ensure we will give you time, and we will stop the interview if you feel to do so. And you are free to decline to answer any questions and to withdraw or stop the interview at any time without any negative consequences to you or your family now or in the future. We will refer you or your family member to the mental health support services available in the site if you wish so.

**Do I have to let my household participate?**

The participation in this survey is voluntary. If you do not agree to participate, we will not ask you any question and we move on with the other households that have been selected. Moreover, even if you agree, you can refuse to answer to some or all questions that will be asked or to stop the interview at any time.

**What are the risks and benefits of participation?**

As with any collection of data, there is always a risk of loss of confidentiality of personal data. However, getting asked personal questions or giving out personal information may be embarrassing or upsetting for you. You can refuse to answer certain questions without any consequences. There is also a risk related to the loss of confidentiality. To decrease this risk, we will work with local personnel who will be meticulously trained in interview techniques and study related documents will be kept in secured in a locked cabinet accessible by the study team only. Neither you nor your family will receive any direct benefit such as food or payment as a reward for participating. However, if someone is ill in your household, he/she will be referred to of our health centres which will be free.

**How will privacy and confidentiality be assured?**

Information collected will be kept confidential and used only for the purpose of the survey. This means we will not ask you for the surname your household, nor the name of your household members. GPS points will be used to identify the randomly selected households the day of the survey but will not be used otherwise. In addition, all documents (both paper and electronic) will be kept in a secure location accessible only to team members. The survey site locations may be used in anonymised analyses in the survey report. The interview will be conducted in the participant household with no interruption to ensure privacy.

After the survey is completed, all paper documents will be archived in a secure room at MSF in Kano state for 5 years and then permanently destroyed. Electronic data will be stored on a secure MSF-OCP server in the European Union. After 5 years the electronic database and all electronic survey data will be permanently deleted.

**How will the results be used?**

Once we have the results of this survey, we will organize meetings with all communities that participated in the research, to inform them of the results. MSF might also use the results of this survey to advocate on behalf of the population at regional, national, or international level to raise awareness of the situation in your community and scientific articles and publications will be also considered as a means of wider dissemination.

Do you have any questions? Please feel free to ask us, we are happy to answer.

In case you have any questions you are free to contact the survey supervisor or the principal investigator whose contacts are indicated below:

Suleiman Hudu at [telephone number removed]

Jonathan Polonsky at [email address removed]

If you have any questions or need further information, you are free to contact the

- MSF office at No. 222 Shehu Kazaure Street, Hotoro GRA, Kano state, [telephone number removed], or
- you may also contact our Data protection officer in case you would like to withdraw your data from the study at
- Kano State Ministry of Health, Post Office Road, Kano state, [telephone number removed]

**Interviewer declaration:** I explained the purpose of the study to the participant and answered all his/her questions. To the best of my knowledge, he/she understands the purpose, procedures, risks and benefits of this study and voluntarily accepts to participate in the study.

**Interviewer name**: _________________________ **Interviewer signature**: _________________________

Date: d|__|__| m|__|__| y|__|__|__|__|

#### Appendix 7: Household survey questionnaire

| **Variable** | **Values** | **Comments** |
| --- | --- | --- |
| **CORE QUESTIONS** | | |
| Team ID | Team ID number |  |
| Date of interview | dd/mm/yyyy |  |
| Cluster ID | Numeric |  |
| Household ID | Numeric | Three-digit number |
| Written informed consent given by the head of the household | Y = Yes N = No |  |
| **HOUSEHOLD QUESTIONS** | | |
| GPS coordinates of household | Lon & Lat (and precision of point) |  |
| *Socio-economic status* |  |  |
| Socioeconomic status:  Household Effects | 0 = None of the following  1 = Radio  2 = Television  3 = Mobile Phone  4 = Computer  5 = Refrigerator  6 = Table  7 = Sofa  8 = Generator  9 = Fan  99 = DNK/NR | Multiple answers possible |
| Socioeconomic status:  Means of Transport | 0 = None of the following  1 = Bicycle  2 = Motorcycle/scooter  3 = Car/truck  4 = Keke napep  99 = DNK/NR | Multiple answers possible |
| Socioeconomic status:  Ownership of agricultural land | Y = Yes  N = No |  |
| Socioeconomic status:  Ownership of farm animals | 0 = None of the following  1 = Cows/bulls/other cattle  2 = Horses/Donkeys  3 = Goats/sheep  4 = Pigs  5 = Chickens/other poultry  6 = Camels  99 = DNK/NR | Multiple answers possible |
| Highest level of education of head of household | 0 = No education  1 = Some primary  2 = Completed primary  3 = Some secondary  4 = Completed secondary  5 = More than secondary |  |
| Highest level of education of female head of household (if there is a male head of household) | 0 = No education  1 = Some primary  2 = Completed primary  3 = Some secondary  4 = Completed secondary  5 = More than secondary |  |
| Main drinking water source | 1 = Surface  2 = Unimproved  3 = Improved  99 = DNK/NR | Surface water (rivers, lakes)  Unimproved water source (Unprotected dug well or unprotected spring)  Improved source (piped water, boreholes, tubewells, protected dug wells, protected springs, rainwater, packaged or delivered water) |
| Main type of toilet used by household members | 1 = Open defecation; 2 = Any unimproved; 3 = Improved 99 = DNK/NR | Open defecation (field, forest, nature)  Any unimproved facility (latrine without slab, bucket)  Any improved facility (toilets, pit latrines with slabs) |
| Handwashing facility with water & soap present at household | 0 = No  1 = Yes  99 = Unable to verify | Check in household |
| *Attitudes to vaccination* |  |  |
| Do you believe that vaccines can protect children from serious diseases? | 0 = No  1 = Yes |  |
| Do you think that most parents like you have their children vaccinated with all the recommended vaccines? | 0 = No  1 = Yes |  |
| Have you ever been reluctant or hesitated to get a vaccination for your child? | 0 = No  1 = Yes |  |
| Have you ever refused a vaccination for your child? | 0 = No  1 = Yes |  |
| If yes to questions 3 or 4, check which ones: |  |  |
| *Diphtheria vaccine* | 1 = Hesitated  2 = Refused |  |
| *DPT3 vaccine* | 1 = Hesitated  2 = Refused |  |
| *Polio vaccine* | 1 = Hesitated  2 = Refused |  |
| *Measles vaccine* | 1 = Hesitated  2 = Refused |  |
| *Meningococcal vaccine* | 1 = Hesitated  2 = Refused |  |
| *“Pentavalent” or other combination infant vaccine* | 1 = Hesitated  2 = Refused |  |
| *BCG vaccine* | 1 = Hesitated  2 = Refused |  |
| Reason for hesitation/refusal | 1 = Did not think it was needed  2 = Did not know where to get vaccination  3 = Did not know where to get good/reliable information  4 = Heard or read negative media  5 = Did not think the vaccine was effective  6 = Did not think the vaccine was safe/concerned about side effects  7 = Someone else told me that the vaccine was not safe  8 = Had a bad experience with previous vaccinator/health clinic  9 = Had a bad experience or reaction with previous vaccination  10 = Fear of needles  11 = Not possible to leave other work  12 = Religious reasons  13 = Refusal of male head of household  14 = Belief it can lead to infertility/sterilisation  15 = Other beliefs/traditional medicine  16 = Other (please specify): |  |
| Has distance ever prevented you from getting your child immunized? | 0 = No  1 = Yes |  |
| Has timing of clinic ever prevented you from getting your child immunized? | 0 = No  1 = Yes |  |
| Have costs ever prevented you from getting your child immunized? | 0 = No  1 = Yes |  |
| Are there other pressures that prevent you from getting your child immunized on time? | 0 = No  1 = Yes |  |
| Are there any reasons why children should not be vaccinated? | 0 = No  1 = Yes |  |
| Do you think it is difficult for some groups to get vaccinations? | 0 = No  1 = Yes |  |
| If yes, reason why: | 1 = They choose not to vaccinate  2 = They do not feel welcome at the health service  3 = Health services do not reach them |  |
| Have you ever received or heard negative information about vaccinations? | 0 = No  1 = Yes (specify) |  |
| If yes, did you still take your child to get vaccinated after? | 0 = No  1 = Yes |  |
| Which leaders in your community support vaccines for children? | 1 = Religious  2 = Political  3 = Teachers  4 = Health care workers 5 = Other (specify) | Multiple answers possible |
| Was your household visited by an MSF health promotion worker who provided information about diphtheria disease, treatment, and vaccination? | 0 = No  1 = Yes  99 = Do not recall |  |
| If yes, did this change how likely you would be to get vaccinations any children in your household? | 0 = No impact  1 = More likely  2 = Less likely  3 = Not relevant (no children in HH) |  |
| How many people are currently living in the household? | Numeric | Used to populate questionnaire (one for each ) |
| **INDIVIDUAL QUESTIONS (For each household member)** | | |
| *Household demography & mortality* |  |  |
| Household member order | Numeric |  |
| Sex | M = Male  F = Female |  |
| Age | Numeric (months/years) |  |
| Arrived during recall period? | 0 = No  1 = Yes |  |
| Date of arrival | (dd/mm/yyyy) |  |
| Left during recall period | 0 = No  1 = Yes |  |
| Date of departure | (dd/mm/yyyy) |  |
| Born during recall period? | 0 = No  1 = Yes |  |
| Date of birth or age in days | (dd/mm/yyyy)  Numeric (days) |  |
| Died during recall period | 0 = No  1 = Yes |  |
| If died, main cause of death? | 1 = Diphtheria  2 = Other respiratory infection  3 = Diarrhoea  4 = Malaria/Fever  5 = Malnutrition  6 = Measles  7 = Meningitis  8 = During pregnancy  9 = During/after delivery (<1 month)  10 = Trauma/accident  11 = Violence  12 = Other (specify)  99 = DNK |  |
| If diphtheria, was there a diagnosis by a health professional? | 0 = No  1 = Yes |  |
| If diphtheria, approximate date of disease onset? | 0 = No  1 = Yes |  |
| If died, place of death | 1 = At current location  2 = At the health centre  3 = At the hospital  4 = At DTC  5 = Other (specify)  99 = Don’t know |  |
| If died, did s/he seek health care in the 2 weeks before dying? | 0 = No  1 = Yes  99 = DNK |  |
| Place health care sought? | 1 = Home  2 = Health centre  3 = Diphtheria Treatment Centre (*specify*)  4 = Private hospital *(specify)*  5 = Public hospital (*specify*)  6 = Traditional healer  7 = Patient Medicine Vendor/Pharmacy  8 = Other place *(specify)*  99 = DNK |  |
| If not at a health facility, what was the main reason for not seeking care in a health structure? | 1 = No money/consultation too expensive  2 = Too sick to seek care  3 = Not sick enough to seek care 4 = Health facility too far away 5 = Went to traditional healer  6 = Did not have time/too busy to go 7 = No confidence in health system  8 = Security problem 9 = Care was refused at facility  10 = Refusal of male head of household 11 = Died suddenly before visit could be arranged  12 = Other reason *(specify)* 99 = DNK |  |
| *Diphtheria vaccination status* |  |  |
| Have you ever received diphtheria vaccination? | 0 = No  1 = Yes |  |
| If yes: Number of doses according to vaccination card | 0 = No card  1 = Yes (1 dose)  2 = Yes (2 doses)  3 = Yes (3 or more doses) |  |
| If card:  Date of vaccination (most recent dose) | (dd/mm/yyyy) |  |
| If no card:  Approximate vaccination date | (MM YYYY) |  |
| Is this individual aged 15 or under? | 0 = No  1 = Yes |  |
| If yes, did they receive the vaccine provided during the campaign? | 0 = No  1 = Yes (card observed)  2 = Yes (card not observed) | Specify months of campaign in question |
| If no, reason why not vaccinated | 1 = Did not think it was needed  2 = Did not know where to get vaccination  3 = Did not know where to get good/reliable information  4 = Heard or read negative media  5 = Did not think the vaccine was effective  6 = Did not think the vaccine was safe/concerned about side effects  7 = Someone else told me that the vaccine was not safe  8 = Had a bad experience with previous vaccinator/health clinic  9 = Had a bad experience or reaction with previous vaccination  10 = Fear of needles  11 = Not possible to leave other work  12 = Religious reasons  13 = Refusal of male head of household  14 = Belief it can lead to infertility  15 = Already vaccinated against diphtheria  16 = Other beliefs/traditional medicine  17 = Other (please specify): | Trying to learn about barriers to vaccination |
| *Diphtheria history* |  |  |
| Did this individual have suspected diphtheria during the recall period | 0 = No 1 = Yes  99 = DNK | Clear case definitions necessary |
| If yes, what was the outcome? | 1 = Recovered  2 = Still unwell (complications)  3 = Died |  |
| If still unwell, what are the long-term health problems? | 1 = Myocarditis/heart failure  2 = Neuritis  3 = Kidney failure  3 = Otitis media  4 = Problems breathing  5 = Malnutrition  6 = Other (specify) | Check all that apply  Will need community language for these, and to provide examples to guide the responders and data collectors, e.g. hoarseness of voice, regurgitation of fluid/food, weakness of limbs… |
| If yes, approximate date of disease onset? | (mm/yy) | Event calendar to help |
| If yes, were they vaccinated prior to disease onset? | 0 = No  1 = Yes (any doses)  99 = DNK | Asking again for extra certainty |
| If yes, did they provide care to another diphtheria patient within the household prior to developing symptoms? | 0 = No 1 = Yes  99 = DNK |  |
| If yes, what were your symptoms? | 1 = Whitish/greyish layer of the tongue  2 = Swollen neck  3 = Fever  4 = Sore throat  5 = Difficulty swallowing  6 = Difficulty breathing  7 = Drooling saliva  8 = Nasal regurgitation  9 = Nasal or other bleeding  10 = Malaise/Lethargy  11 = Loss of appetite  12 = Hoarseness  99 = None of the above | Check all that apply  Will need community language for these |
| If yes, was there a diagnosis by a health professional? | 0 = No  1 = Yes  99 = DNK |  |
| If yes, did they seek healthcare? | 0 = No 1 = Yes  99 = DNK |  |
| If yes, what treatment did they receive? | 0 = None  1 = Antibiotic only  2 = Antitoxin only  3 = Antibiotic and antitoxin  4 = Other/traditional treatment  99 = DNK |  |
| If yes, place health care sought? | 1 = Home  2 = Health centre  3 = Diphtheria Treatment Centre (*specify*)  4 = Private hospital *(specify)*  5 = Public hospital (*specify*)  6 = Traditional healer  7 = Patient Medicine Vendor/Pharmacy  8 = Other place *(specify)*  99 = DNK | Select all that apply, in order of visit |
| If yes, after seeking healthcare, where were they finally treated? | 1 = Health facility (specify)  2 = Home  99 = DNK | This is to distinguish between those receiving DTC-based and home-based care |
| Did the treatment lead to an improvement in the condition? | 1 = Strongly disagree  2 = Disagree  3 = No opinion  4 = Agree  5 = Strongly agree  99 = DNK |  |
| If quite/strongly negative, please explain | Free text |  |
| How acceptable was the treatment to you? | 1 = Completely unacceptable  2 = Unacceptable  3 = No opinion  4 = Acceptable  5 = Completely acceptable  99 = DNK |  |
| If quite/strongly negative, please explain | Free text |  |
| How do you feel about the quality of treatment you received? | 1 = Strongly positive  2 = Quite positive  3 = Neutral  4 = Quite negative (specify)  5 = Strongly negative (specify) |  |
| If quite/strongly negative, please explain | Free text |  |
| If at home, how do you feel about being treated at home rather than in a health facility? | 1 = Strongly positive  2 = Quite positive  3 = Neutral  4 = Quite negative (specify)  5 = Strongly negative (specify) |  |
| If quite/strongly negative, please explain | Free text |  |
| If yes, who treated them? | 1 = MSF  2 = Other NGO (e.g. Alima/Save The Children) 3 = Government health facilities not supported by NGOs  99 = DNK |  |
| If yes, delay between disease onset and treatment beginning? | Numeric (days) | This refers to diphtheria treatment by one of the providers above (NGO/govt) |
| If no, main reason for not seeking care in a health structure? | 1 = No money/consultation too expensive  2 = Too sick to seek care  3 = Not sick enough to seek care 4 = Health facility too far away 5 = Went to traditional healer  6 = Did not have time/too busy to go 7 = No confidence in health system  8 = Security problem 9 = Care was refused at facility  10 = Refusal of male head of household 11 = Died suddenly before visit could be arranged  12 = Other reason *(specify)* 99 = DNK |  |

#### Appendix 8: Retrospective cohort study questionnaire

| **CORE QUESTIONS** |  |  |
| --- | --- | --- |
| **Variable** | **Values** | **Comments** |
| Team ID | Team ID number |  |
| Date of interview | dd/mm/yyyy |  |
| Patient ID | Numeric |  |
| Written informed consent given by the respondent | Y = Yes N = No |  |
| Address of household | Free text | For comparison with records to assess biases in residential location recording |
| GPS coordinates of household | Lon & Lat (and precision of point) |  |
| How many people are currently living in the household? | Numeric |  |
| *Diphtheria vaccination status* |  |  |
| Have you ever received diphtheria vaccination? | 0 = No  1 = Yes |  |
| If yes: Number of doses according to vaccination card | 0 = No card  1 = Yes (1 dose)  2 = Yes (2 doses)  3 = Yes (3 or more doses) |  |
| If card:  Date of vaccination (most recent dose) | (dd/mm/yyyy) |  |
| If no card:  Approximate vaccination date | (MM YYYY) |  |
| Is this individual aged 15 or under? | 0 = No  1 = Yes | Options available will include known periods of campaigns |
| If yes, did they receive the vaccine provided during the campaign? | 0 = No  1 = Yes (card observed)  2 = Yes (card not observed) | Specify months of campaign in question |
| If no, reason why not vaccinated | 1 = Did not think it was needed  2 = Did not know where to get vaccination  3 = Did not know where to get good/reliable information  4 = Heard or read negative media  5 = Did not think the vaccine was effective  6 = Did not think the vaccine was safe/concerned about side effects  7 = Someone else told me that the vaccine was not safe  8 = Had a bad experience with previous vaccinator/health clinic  9 = Had a bad experience or reaction with previous vaccination  10 = Fear of needles  11 = Not possible to leave other work  12 = Religious reasons  13 = Refusal of male head of household  14 = Belief it can lead to infertility  15 = Already vaccinated against diphtheria  16 = Other beliefs/traditional medicine  17 = Other (please specify): |  |
| Was your household visited by an MSF health promotion worker who provided information about diphtheria and vaccination? | 0 = No  1 = Yes  99 = Do not recall |  |
| If yes, did this change how likely you would be to get vaccinations for yourself and your family? | 0 = No impact  1 = More likely  2 = Less likely |  |
| *Diphtheria history* |  |  |
| What were your symptoms? | 1 = Whitish/greyish layer of the tongue  2 = Swollen neck  3 = Fever  4 = Sore throat  5 = Difficulty swallowing  6 = Difficulty breathing  7 = Drooling saliva  8 = Nasal regurgitation  9 = Nasal blood  10 = Malaise/Lethargy  11 = Loss of appetite  12 = Hoarseness  99 = None of the above | Check all that apply  Will need community language for these |
| What was the outcome of illness? | 1 = Recovered  2 = Still unwell  3 = Died |  |
| If still unwell, what are the long-term health problems? | 1 = Myocarditis/heart failure  2 = Neuritis  3 = Kidney failure  3 = Otitis media  4 = Problems breathing  5 = Malnutrition  6 = Other (specify) | Check all that apply  Will need community language for these |
| Where was the first place you sought healthcare? | 1 = Health centre (*specify*)  2 = Diphtheria Treatment Centre (*specify*)  3 = Hospital *(specify)*  4 = Traditional healer  5 = Pharmacy  6 = Other place *(specify)*  99 = DNK |  |
| If not at a health facility, what was the main reason for not seeking care in a health structure? | 1 = No money/consultation too expensive  2 = Too sick to seek care  3 = Not sick enough to seek care 4 = Health facility too far away 5 = Went to traditional healer  6 = Did not have time/too busy to go 7 = No confidence in health system  8 = Security problem 9 = Care was refused at facility  10 = Other reason *(specify)*  99 = DNK |  |
| What was the delay between disease onset and treatment beginning with MSF? | Numeric (days) |  |
| Where were you treated by MSF? | 1 = At health facility  2 = At home |  |
| What treatment did you receive? | 0 = None  1 = Antibiotic only  2 = Antitoxin only  3 = Antibiotic and antitoxin  4 = Other/traditional treatment  99 = DNK |  |
| Did your $home-based/$hospital-based treatment lead to an improvement in the condition? | 1 = Strongly disagree  2 = Disagree  3 = No opinion  4 = Agree  5 = Strongly agree  99 = DNK |  |
| If quite/strongly negative, please explain | Free text |  |
| How acceptable was the $home-based/$hospital-based treatment to you? | 1 = Completely unacceptable  2 = Unacceptable  3 = No opinion  4 = Acceptable  5 = Completely acceptable  99 = DNK |  |
| If quite/strongly negative, please explain | Free text |  |
| How do you feel about the quality of treatment you received? | 1 = Strongly positive  2 = Quite positive  3 = Neutral  4 = Quite negative (specify)  5 = Strongly negative (specify) |  |
| If quite/strongly negative, please explain | Free text |  |
| If at home, how do you feel about being treated at home rather than in a health facility? | 1 = Strongly positive  2 = Quite positive  3 = Neutral  4 = Quite negative (specify)  5 = Strongly negative (specify) |  |
| If quite/strongly negative, please explain | Free text |  |
| Did anyone else in your household show signs of diphtheria prior to you developing symptoms? | 0 = No 1 = Yes  99 = DNK |  |
| If yes, what was the delay between their disease beginning and yours beginning? | Numeric (days/weeks) |  |
| Did anyone else in your household show signs of diphtheria after you developed symptoms? | 0 = No 1 = Yes  99 = DNK |  |
| If yes, how many people? | Numeric |  |
| If yes, what was the delay between the beginning of your disease and the next person to fall ill? | Numeric (days/weeks) |  |
| *For other household members* |  |  |
| Household member order | Numeric |  |
| Sex | M = Male  F = Female |  |
| Age | Numeric (months/years) |  |
| Did this person ever receive diphtheria vaccination? | 0 = No  1 = Yes |  |
| If yes: Number of doses according to vaccination card | 0 = No card  1 = Yes (1 dose)  2 = Yes (2 doses)  3 = Yes (3 or more doses) |  |
| If card:  Date of vaccination (most recent dose) | (dd/mm/yyyy) |  |
| If no card:  Approximate vaccination date | (MM YYYY) |  |
| Did this individual have suspected diphtheria during the recall period | 0 = No 1 = Yes  99 = DNK | Clear case definitions necessary |
| If yes, did they develop symptoms before or after the subject of this investigation? | 1 = Before  2 = After  99 = Do not recall |  |
| If yes, what was the outcome? | 1 = Recovered  2 = Still unwell (complications)  3 = Died |  |
| If yes, approximate date of disease onset? | (mm/yy) | Event calendar to help |
| If yes, were they vaccinated prior to disease onset? | 0 = No  1 = Yes (any doses)  99 = DNK | Asking again for extra certainty |
| If yes, did they provide care to another diphtheria patient within the household prior to developing symptoms? | 0 = No 1 = Yes  99 = DNK |  |
| If yes, what were their symptoms? | 1 = Whitish/greyish layer of the tongue  2 = Swollen neck  3 = Fever  4 = Sore throat  5 = Difficulty swallowing  6 = Difficulty breathing  7 = Drooling saliva  8 = Nasal regurgitation  9 = Nasal or other bleeding  10 = Malaise/Lethargy  11 = Loss of appetite  12 = Hoarseness  99 = None of the above | Check all that apply  Will need community language for these |
| If yes, was there a diagnosis by a health professional? | 0 = No  1 = Yes  99 = DNK |  |
| If yes, did they seek healthcare? | 0 = No 1 = Yes  99 = DNK |  |
| If yes, what treatment did they receive? | 0 = None  1 = Antibiotic only  2 = Antitoxin only  3 = Antibiotic and antitoxin  4 = Other/traditional treatment  99 = DNK |  |
| If yes, place health care sought? | 1 = Home  2 = Health centre  3 = Diphtheria Treatment Centre (*specify*)  4 = Private hospital *(specify)*  5 = Public hospital (*specify*)  6 = Traditional healer  7 = Patient Medicine Vendor/Pharmacy  8 = Other place *(specify)*  99 = DNK | Select all that apply, in order of visit |
| If yes, after seeking healthcare, where were they finally treated? | 1 = Health facility (specify)  2 = Home  99 = DNK | This is to distinguish between those receiving DTC-based and home-based care |
| If yes, who treated them? | 1 = MSF  2 = Other NGO (e.g. Alima/Save The Children) 3 = Government health facilities not supported by NGOs  99 = DNK |  |
| If yes, delay between disease onset and treatment beginning? | Numeric (days) | This refers to diphtheria treatment by one of the providers above (NGO/govt) |
| If no, main reason for not seeking care in a health structure? | 1 = No money/consultation too expensive  2 = Too sick to seek care  3 = Not sick enough to seek care 4 = Health facility too far away 5 = Went to traditional healer  6 = Did not have time/too busy to go 7 = No confidence in health system  8 = Security problem 9 = Care was refused at facility  10 = Refusal of male head of household 11 = Died suddenly before visit could be arranged  12 = Other reason *(specify)* 99 = DNK |  |
